# Sex Differences in the Risk of Bronchopulmonary Dysplasia and Pulmonary Hypertension: A Bayesian Meta-Analysis

**DOI:** 10.1101/2025.01.22.25320939

**Authors:** Elke van Westering-Kroon, Tamara M. Hundscheid, Karen Van Mechelen, František Bartoš, Steven H. Abman, Eduardo Villamor

## Abstract

**Background:** Bronchopulmonary dysplasia (BPD) is generally considered to be more frequent in males than in females. We conducted a Bayesian model-averaged (BMA) meta-analysis of studies addressing sex differences in the risk of developing different severities of BPD and BPD-associated pulmonary hypertension (BPD-PH).

**Methods:** We used BMA to calculate Bayes factors (BFs). The BF_10_ is the ratio of the probability of the data under the alternative hypothesis (presence of sex differences) over the probability of the data under the null hypothesis (absence of sex differences). BPD was classified as BPD28 (supplemental oxygen at or during 28 days), BPD36 (oxygen at 36 weeks postmenstrual age), mild, moderate, and severe BPD.

**Results:** We included 222 studies (541,826 infants). The BMA analysis showed extreme evidence in favor of a male disadvantage in BPD28 (BF_10_>10^5^), BPD36 (BF_10_>10^21^), and severe BPD (BF_10_=87.55), but not in mild BPD (BF_10_=0.28), or BPD-PH (BF_10_=0.54). The evidence for a male disadvantage in BPD decreased as the gestational age of the cohort decreased.

**Conclusions:** We confirmed the presence of a male disadvantage in moderate-to-severe BPD, but not in less severe forms of BPD or in BPD-PH. The male disadvantage in BPD is much less apparent in the more immature infants.

**Impact:** This Bayesian meta-analysis confirms that the risk of developing moderate to severe bronchopulmonary dysplasia (BPD) is approximately 20% higher in males than in females.

Sex differences in BPD decrease with decreasing gestational age, are heterogeneous across geographic and sociodemographic settings, and have remained persistently stable over time.

There is no evidence supporting sex differences in pulmonary hypertension associated with BPD.

An important step in the process of individualizing the approach to BPD may be to consider the sex of the infant, as this information can be used to personalize care and potentially improve outcomes.

## Introduction

Bronchopulmonary dysplasia (BPD), the chronic lung disease of prematurity, is a common complication of preterm birth and a major contributor to infant mortality and morbidity in both the short and long term. ^1 2^ Low gestational age (GA) is the main risk factor for developing BPD.^1 2^ However, the etiopathogenesis of BPD is multifactorial with numerous pre- and postnatal factors that may contribute to its development.^1 2^ The incidence of BPD continues to increase because advances in neonatal and perinatal care have led to improved survival of the smallest newborn infants.^1^ Pulmonary hypertension (PH) complicates the course of BPD in 17–37% of preterm infants.^3 4^ BPD-associated PH (BPD-PH) is linked to higher mortality rates and increased morbidity in survivors.^3 4^

A widely accepted concept in perinatal medicine is the so-called ‘male disadvantage of prematurity’. This refers to the observation that mortality and major complications of prematurity are more frequent in males than in females.^5-7^ A large body of evidence supports BPD as part of this male disadvantage of prematurity.^6 7^ However, it has not been determined whether sex differences in BPD rates are present at all levels of BPD severity and across all geographic settings. It is also unknown whether the male disadvantage has been altered by epidemiological changes in BPD due to increased survival of infants with the lowest GAs.^7^ Therefore, our objective was to perform a systematic review and meta-analysis of studies reporting sex-specific BPD rates for different degrees of severity of the condition. Additionally, we aimed to investigate whether sex differences affected BPD-PH. A further aim was to investigate, through subgroup analyses, whether sex differences in BPD and BPD-PH depend on GA and are consistent across different geographical and socio-demographic settings. Finally, we used meta-regression to examine whether sex differences in BPD have evolved over time.

Instead of the more common frequentist statistics with *p*-values and confidence intervals, we used a Bayesian approach to meta-analysis.^8^ An important limitation of frequentist null hypothesis (H_0_) significance testing is that failing to reject H_0_ when the *p*-value is below a predetermined threshold (usually 0.05) or the confidence interval intersects the line of no effect, does not mean evidence for H_0_. Conversely, the rejection of H_0_ when the *p*-value is above the threshold does not necessarily mean that we have found evidence in support of the alternative hypothesis (H_1_).^8^ Using a Bayesian approach, the strength of the evidence can be tested in terms of both H_1_ (presence of sex differences) and H_0_ (absence of sex differences). ^8^

## Methods

The methodology of this study is based on that of earlier studies by our group on sex differences and risk factors for outcomes of prematurity.^4 5^ The study was performed and reported according to the preferred reporting items for systematic reviews and meta-analyses and meta-analysis of observational studies in epidemiology guidelines. The review protocol was registered in the PROSPERO international register of systematic reviews (CRD42018095509). The research question was “Do preterm boys have a different risk of developing BPD and BPD-PH than preterm girls?”

### Sources and search strategy

A comprehensive literature search was undertaken using the PubMed and Embase databases. The search strategy is detailed in Supplementary Table 1. No language limit was applied. The literature search was updated up to September 2023.

### Study selection and definitions

Studies were included if they had a prospective or retrospective cohort design, examined preterm infants (GA < 37 weeks) and reported primary data that could be used to measure the association between infant sex and rate of BPD. Studies that exclusively included late preterm infants (GA ≥ 34 weeks) or combined preterm and term infants were excluded. To identify relevant studies, two reviewers (EvW-K and TMH) independently screened the results of the searches and applied inclusion criteria using a structured form. Discrepancies were identified and resolved through discussion or in consultation with the other researchers.

The definition and categorization of BPD was based on the combination of the classic criteria of 28 days postnatal life and 36 weeks postmenstrual age (PMA).^9^ BPD28 was defined as need of oxygen or respiratory support during 28 days or at postnatal day 28. BPD36 was defined as need of oxygen or respiratory support at 36 weeks PMA. BPD severity was categorized according to the support required at 36 weeks PMA. Infants in room air at 36 weeks PMA were categorized as having mild BPD, those requiring 22 to <30% oxygen as having moderate BPD, and those requiring ≥30% oxygen and/or positive pressure as having severe BPD.^9^ BPD-PH was defined by any echocardiographic criteria, provided the evaluation was performed at postnatal age >28 days.^10^

### Data extraction and assessment of risk of bias

Two investigators (EvW-K, and TMH) extracted data on the study design, demographics, and rate of BPD. A second group of investigators (KvM and EV) checked the data extraction for completeness and accuracy. Risk of bias was assessed using the Newcastle1.Ottawa scale (NOS) for cohort studies^11^ and the Quality in Prognosis Studies (QUIPS) tool, with the modifications proposed by Stallings et al. for the study of sex as a prognostic factor.^12^ Details of the tools are provided in the Supplementary Methods.

### Bayesian model-averaged meta-analysis

Values of log risk ratio (logRR) or log odds ratio (logOR) and the corresponding standard error of each individual study were calculated using comprehensive meta-analysis (CMA) V4.0 software (Biostat Inc., Englewood, NJ). The results were further pooled and analyzed by a Bayesian model-averaged (BMA) meta-analysis.^7 8^ We performed the BMA in JASP, which utilizes the metaBMA R package.^13 14^ BMA employs Bayes factors (BFs) and Bayesian model-averaging to evaluate the likelihood of the data under the combination of models assuming the presence vs. the absence of the meta-analytic effect and heterogeneity.^8^ The BF_10_ is the ratio of the probability of the data under H_1_ over the probability of the data under H_0_. The BF_10_ was interpreted using the evidence categories suggested by Lee & Wagenmakers^15^ (Figure 1 and Supplementary Methods). The BF_rf_ is the ratio of the probability of the data under the random effects model over the probability of the data under the fixed effect model. We used the neonatal-specific empirical prior distributions based on the Cochrane Database of Systematics Review: logRR ∼ Student-t(µ = 0, σ = 0.18, ν = 3), tau(logRR) ∼ Inverse-Gamma(k = 1.89, θ = 0.30); logOR∼ Student-t(µ = 0, σ = 0.29, ν = 3), tau(logOR) ∼ Inverse-Gamma(k = 1.80, θ = 0.42).^16^ We used robust Bayesian meta-analysis (RoBMA) to assess the robustness of the results to the potential presence of publication bias, which was expressed as BF_bias_ (Supplementary Methods).^17^

### Subgroup analysis and meta-regression

In order to include the maximum number of cohorts, our inclusion criterion for GA was broad (GA < 37 weeks). However, BPD is a condition that almost exclusively affects extremely preterm infants (GA < 28 weeks). To ascertain whether sex differences in BPD remained consistent across the various GA subgroups, we conducted an analysis in which the studies were classified into four subgroups: mean or median GA ≥ 27 weeks, <27 weeks, ≤25 weeks, and ≤23 weeks. In instances where a study provided data specific to a GA subgroup, these data were incorporated into the corresponding subgroup analysis. Since RRs depend on the baseline risk, ^18^ and the baseline risk of BPD is inversely proportional to GA, we also used ORs in the analysis of GA subgroups.

**Figure 1.**
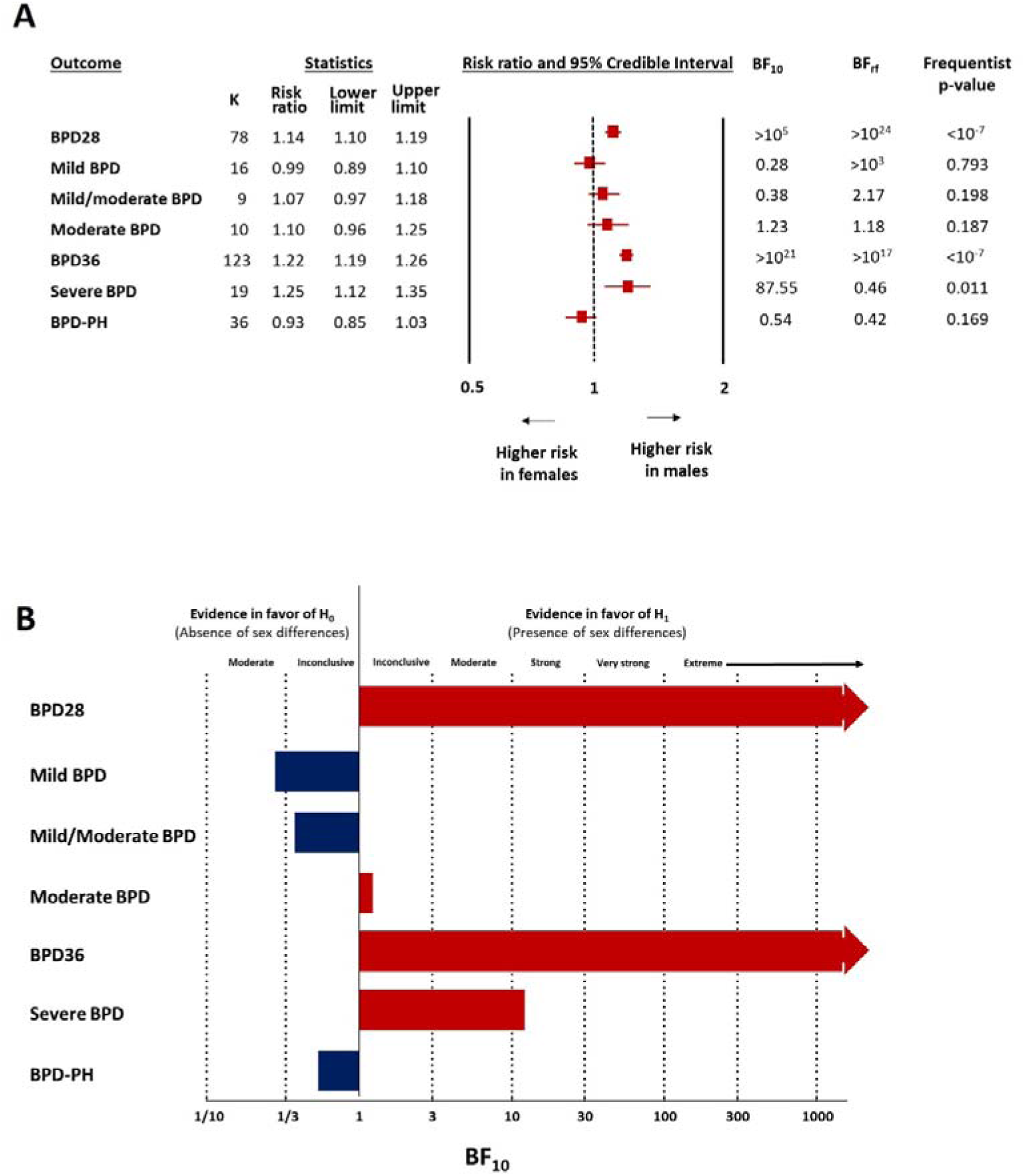
Summary of Bayesian model average meta-analyses on the association between infant sex and bronchopulmonary dysplasia (BPD). **A.** Summary of risk ratios (RRs). RR>1 indicates higher risk of BPD in males. The p-value was obtained using random-effects frequentist meta-analysis. **B.** Summary of Bayes factors (BFs). The BF_10_ is the ratio of the probability of the data under the alternative hypothesis (H_1_) over the probability of the data under the null hypothesis (H_0_).

To examine potential variations in sex differences in BPD across geographic and sociodemographic areas, we conducted a subgroup analysis by (sub)continent and sociodemographic index (SDI) quintile. The SDI is a composite measure of developmental status as it is associated with health (Supplementary Methods). ^19^ For comparisons across SDI quintiles, each country was assigned to a single quintile according to its SDI in 2019.

Finally, to examine potential changes over time in sex differences in BPD, we conducted a frequentist random effects meta-regression (method of moments) plotting the median year of birth of each cohort against the effect size (logRR) of the association between infant sex and BPD. We used the CMA software to perform the meta-regression analysis.

## Results

### Description of studies and risk of bias assessment

The flow diagram of the search process is shown in online supplemental eFigure 1. Of 4722 potentially relevant studies, 222 were included (541,826 infants). Their characteristics are summarized in online supplemental eTable 1. The quality score of each study according to the NOS is depicted in online supplemental eTable 1. All studies received a score of six or higher, indicating a low to moderate risk of bias. Upon the application of the modified QUIPS tool, no evidence of bias by sex was identified in any of the six domains across the studies.

### Bayesian model averaged meta-analysis

Figure 1 and Tables 1-2 summarize the results of the BMA meta-analysis. LogRR was converted to RR for clarity. BMA analysis showed extreme evidence in favor of H_1_ (presence of sex differences) for BPD28 (BF_10_>10^5^, Figure 1, Table 1, online supplemental eFigure 2) and BPD36 (BF_10_>10^21^, Figure 1, Table 1, online supplemental eFigure 3), and very strong evidence in favor of H_1_ for severe BPD (BF_10_=87.55, Figure 1, Table 2, online supplemental eFigure 4). Sex differences in BPD28, BPD36, and severe BPD were in the form of a male disadvantage. In contrast, BMA analysis showed moderate evidence in favor of H_0_ (absence of sex differences) for mild BPD (BF_10_=0.28, Figure 1, online supplemental eFigure 5). Evidence in favor of H_0_ was weak/inconclusive for mild to moderate BPD (BF_10_=0.38, Figure 1, online supplemental eFigure 6), whereas evidence in favor of H_1_ was weak/inconclusive for moderate BPD (BF_10_=1.18, Figure 1, online supplemental eFigure 7). With regard to BPD-PH, the BMA analysis showed weak/inconclusive evidence in favor of H_0_ (BF_10_=0.54, Figure 1, Table 2, online supplemental eFigure 8).

**Figure 2.**
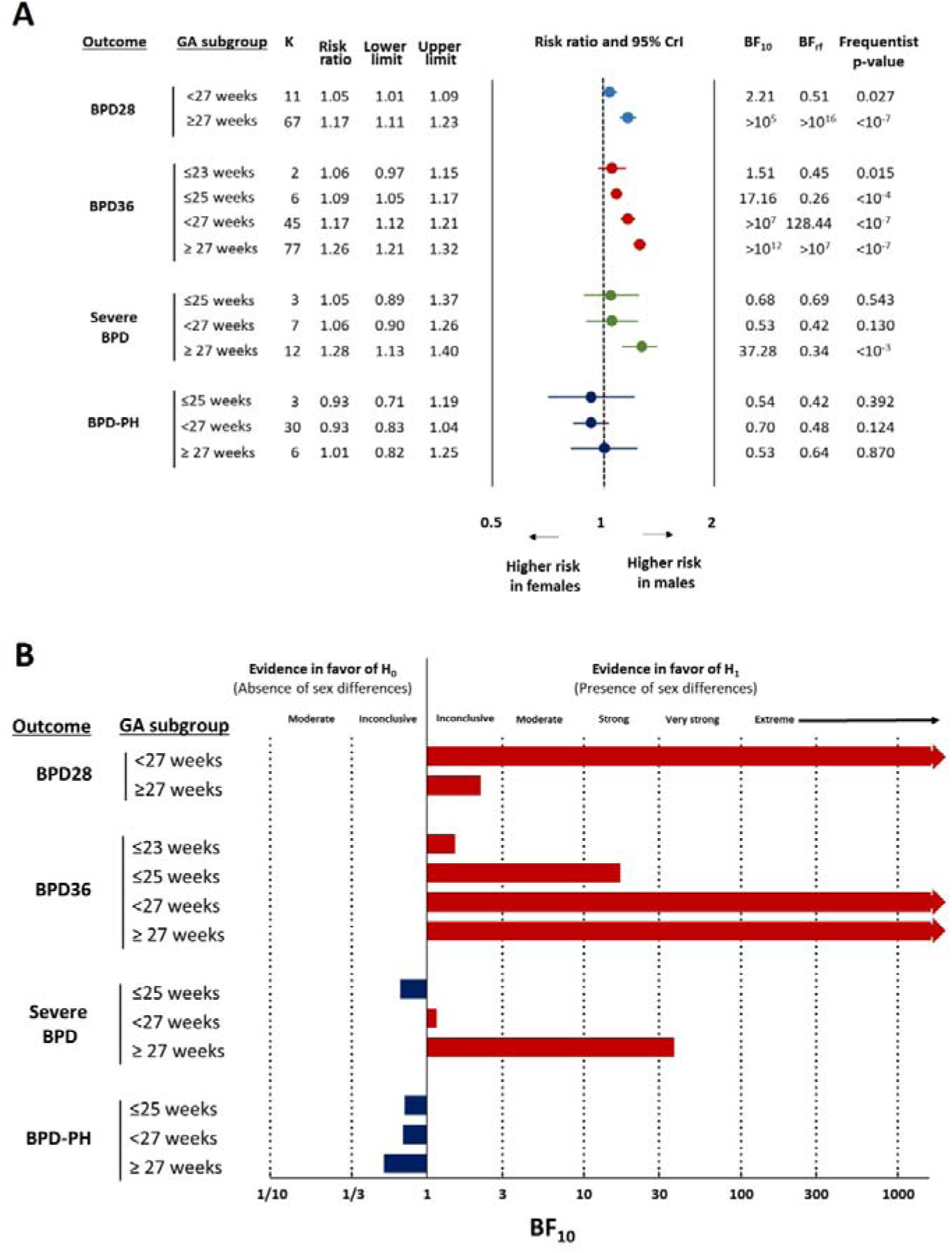
Summary of the subgroup analysis based on the mean or median gestational age (GA) of the cohort. **A.** Summary of risk ratios (RRs). RR>1 indicates higher risk of BPD in males. The p-value was obtained using random-effects frequentist meta-analysis. **B.** Summary of Bayes factors (BFs). The BF_10_ is the ratio of the probability of the data under the alternative hypothesis (H_1_) over the probability of the data under the null hypothesis (H_0_).

**Figure 3.**
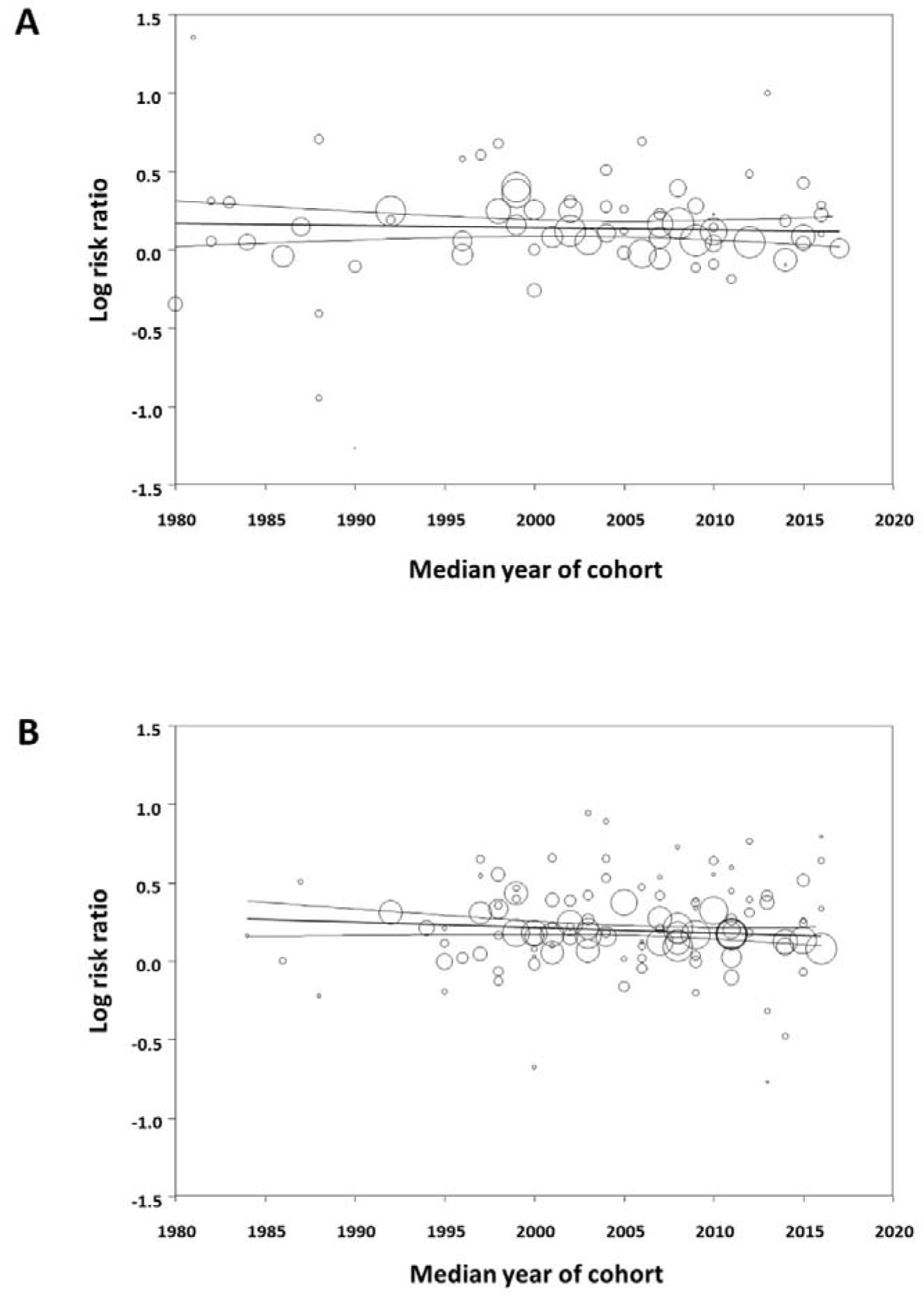
Meta regression. Plot showing the correlation between the association of male sex with BPD28 (A) and BPD36 (B) and the median year of birth of the infants of each cohort. The size of the circles represents the relative weight of each study. R^2^-analog was 0 for both meta-regressions.

**Table 1.** Bayesian model averaged meta-analysis of the association between infant sex and bronchopulmonary dysplasia defined at 28 days of postnatal age (BPD28) or 36 weeks of postmenstrual age (BPD36)

| Outcome | Subgroup |  | K | Averaged effect (RR) | Credible interval |  | BF<sub>10</sub> | P-value frequentist analysis<sup>a</sup> | BF<sub>rf</sub> |
| --- | --- | --- | --- | --- | --- | --- | --- | --- | --- |
|  |  |  |  |  | Lower Limit | Upper Limit |  |  |  |
| BPD28 | All |  | 78 | 1.14 | 1.10 | 1.19 | >10<sup>5</sup> | <10<sup>-7</sup> | >10<sup>24</sup> |
|  | (sub)Continent | Eastern Asia | 16 | 1.07 | 0.92 | 1.25 | 0.52 | 0.010 | >10<sup>24</sup> |
|  |  | Europe | 26 | 1.13 | 1.04 | 1.22 | 10.54 | 0.004 | >10<sup>4</sup> |
|  |  | Latin America | 4 | 1.30 | 1.07 | 1.57 | 14.87 | <10<sup>-4</sup> | 0.70 |
|  |  | North America | 23 | 1.10 | 1.04 | 1.17 | 12.53 | <10<sup>-7</sup> | 766.52 |
|  |  | Oceania | 3 | 1.13 | 0.87 | 1.51 | 0.96 | 0.208 | 0.84 |
|  |  | Western Asia | 6 | 1.23 | 0.96 | 1.51 | 3.29 | 0.020 | 120.43 |
|  | SDI | High | 51 | 1.12 | 1.07 | 1.18 | 869.17 | <10<sup>-7</sup> | >10<sup>12</sup> |
|  |  | High-Middle | 16 | 1.17 | 1.05 | 1.31 | 10.61 | 0.004 | >10<sup>4</sup> |
|  |  | Middle | 10 | 1.18 | 1.02 | 1.38 | 4.70 | 0.015 | 0.78 |
| BPD36 | All |  | 123 | 1.22 | 1.19 | 1.26 | >10<sup>21</sup> | <10<sup>-7</sup> | >10<sup>17</sup> |
|  | (sub)Continent | Eastern Asia | 15 | 1.18 | 1.13 | 1.22 | 322.52 | <10<sup>-7</sup> | 0.11 |
|  |  | Europe | 47 | 1.27 | 1.19 | 1.37 | >10<sup>7</sup> | <10<sup>-7</sup> | >10<sup>10</sup> |
|  |  | Latin America | 3 | 1.44 | 0.95 | 2.19 | 2.45 | 0.089 | 1.51 |
|  |  | North America | 51 | 1.20 | 1.16 | 1.24 | >10<sup>9</sup> | <10<sup>-7</sup> | 19.70 |
|  |  | Oceania | 4 | 1.22 | 0.99 | 1.50 | 1.96 | 0.063 | 8.66 |
|  |  | Western Asia | 2 | 1.48 | 1.38 | 1.59 | 14.76 | <10<sup>-7</sup> | 0.57 |
|  | SDI | High | 100 | 1.19 | 1.16 | 1.23 | >10<sup>16</sup> | <10<sup>-7</sup> | >10<sup>8</sup> |
|  |  | High-Middle | 18 | 1.44 | 1.37 | 1.51 | >10<sup>6</sup> | <10<sup>-7</sup> | 0.13 |
|  |  | Middle | 4 | 1.13 | 0.87 | 1.45 | 0.74 | 0.360 | 0.77 |

*BF* Bayes factor, *BPD2RR* risk ratio (RR>1 indicates higher risk in males), *SDI* sociodemographic index.
<sup>a</sup>Random effects frequentist meta-analysis

**Table 2.** Bayesian model averaged meta-analysis of the association between infant sex and severe bronchopulmonary dysplasia (BPD) and BPD-associated pulmonary hypertension (BPD-PH).

| Outcome | Subgroup |  | K | Averaged effect (RR) | Credible interval |  | BF<sub>10</sub> | P-value frequentist analysis<sup>a</sup> | BF<sub>rf</sub> |
| --- | --- | --- | --- | --- | --- | --- | --- | --- | --- |
|  |  |  |  |  | Lower Limit | Upper Limit |  |  |  |
| Severe BPD | All |  | 19 | 1.25 | 1.12 | 1.35 | 87.55 | <10<sup>-4</sup> | 0.46 |
|  | (sub)Continent | Eastern Asia | 6 | 1.18 | 0.96 | 1.46 | 2.45 | 0.116 | 0.91 |
|  |  | Europe | 5 | 1.34 | 1.00 | 1.82 | 5.61 | 0.050 | 1.72 |
|  |  | North America | 7 | 1.25 | 1.07 | 1.40 | 14.94 | 0.005 | 1.58 |
|  | SDI | High | 13 | 1.21 | 1.07 | 1.38 | 15.09 | 0.002 | 0.75 |
|  |  | High-Middle | 2 | 1.92 | 1.12 | 3.28 | 1.87 | 0.018 | 1.26 |
|  |  | Middle | 2 | 0.52 | 0.14 | 1.87 | 1.01 | 0.329 | 0.89 |
| BPD-PH | All |  | 36 | 0.93 | 0.85 | 1.03 | 0.54 | 0.169 | 0.42 |
|  | (sub)Continent | Eastern Asia | 8 | 0.96 | 0.75 | 1.24 | 0.51 | 0.765 | 0.68 |
|  |  | Europe | 6 | 1.02 | 0.74 | 1.41 | 0.62 | 0.916 | 0.60 |
|  |  | North America | 20 | 0.91 | 0.80 | 1.03 | 0.76 | 0.121 | 0.60 |
|  | SDI | High | 31 | 0.94 | 0.85 | 1.05 | 0.44 | 0.287 | 0.48 |
|  |  | Middle | 3 | 0.85 | 0.65 | 1.11 | 0.86 | 0.227 | 0.71 |

BF Bayes factor, RR risk ratio (RR>1 indicates higher risk in males), SDI sociodemographic index.
<sup>a</sup>Random effects frequentist meta-analysis

BMA analysis showed extreme evidence of heterogeneity (BF_rf_>100) in the BPD28, mild BPD, and BPD36 analyses (Figure 1). Evidence for heterogeneity was inconclusive for the other meta-analyses. Detailed heterogeneity data are provided in online supplemental eTables 2-6. RoBMA showed moderate evidence of publication bias for the mild to moderate BPD meta-analysis (BF_bias_=4.43).

Following adjustment for publication bias, RoBMA showed moderate evidence in favor of H_0_ for this association (BF_10_=0.32). Evidence against or for publication bias was inconclusive for the other analyses (online supplemental eTable 7).

### Subgroup analysis and meta-regression

Subgroup analyses based on the GA of the cohort (mean or median GA>27 weeks, <27 weeks, or ≤25 weeks) were conducted for BPD28, BPD36, severe BPD, and BPD-PH. As shown in Figure 2, the effect size (expressed as logRR) and the strength of evidence (expressed as BF10) for the male disadvantage in BPD were smaller in the subgroups that included cohorts with lower mean or median GA. To evaluate the robustness of the GA subgroup analysis, we conducted it using the OR as a measure of the effect size. The reduction with the GA of the cohort in both the effect size and the strength of the evidence of sex differences was also observable when the effect size was expressed as the OR (online supplemental eTable 8).

Subgroup analyses based on (sub)continent and SDI were conducted for BPD28 (Table 1), BPD36 (Table 1), severe BPD (Table 2) and BPD-PH (Table 2). Regarding BPD28, evidence in favor of H_0_ was weak for Eastern Asia (BF_10_=0.52) and Oceania (BF_10_=0.96), and strong in favor of H_1_ for Europe, Latin America, and North America (Table 1). The evidence in favor of H_1_ for BPD28 was extreme for the high SDI subgroup (BF_10_=869.17), strong for the high-middle SDI subgroup (BF_10_=10.61), and moderate for the middle SDI subgroup (BF_10_=4.70). Regarding BPD36, evidence in favor of H_1_ was weak for Latin America (BF_10_=2.45), and Oceania (BF_10_=1.96). The evidence in favor of H_0_ was weak for the subgroup middle SDI (BF_10_=0.74) (Table 2). Regarding severe BPD, evidence in favor of H_1_ was strong for North-America (BF_10_=14.94), moderate for Europe (BF_10_=5.61), and weak for Eastern Asia (BF_10_=2.45) (Table 2). The evidence in favor of H_1_ for severe BPD was strong for the high SDI subgroup (BF_10_=15.09), but weak for the high-middle (BF_10_=1.87), and middle SDI (BF_10_=1.01) subgroups (Table 2). Regarding BPD-PH, evidence in favor of H_0_ was weak for all the subgroups (Table 2).

Subgroup analysis based on the number of infants included in the study (greater or less than 500) was only conducted for BPD28 and BPD36, as these were the two meta-analyses with the largest number of studies. This subgrouping did not result in substantial changes in effect size or strength of evidence (supplemental eTable 9).

Finally, meta-regression showed that the effect size of the association between sex and BPD28 as well as BPD36 remained stable over time. That is, it did not correlate with the median year of birth of the cohort (Figure 2).

## Discussion

This study represents a comprehensive examination of sexual dimorphism in BPD. Our findings confirm the robust evidence of a male disadvantage in moderate to severe BPD. The risk of developing BPD36 or severe BPD is approximately 20% higher in males than in females. Sex differences in BPD decrease with decreasing GA, are heterogeneous across geographic and sociodemographic settings, and have remained persistently stable over time. Notably, despite the well-documented association between PH and the most severe forms of BPD, ^2 3^ our findings suggest that male disadvantage does not involve BPD-PH. Furthermore, sex differences were not present for mild BPD.

BPD is increasingly recognized as the consequence of a pathological reparative response of the lung of infants born in the late canalicular or early saccular stage of pulmonary development to both prenatal and postnatal stressors. Prenatal insults, such as placental vascular insufficiency or chorioamnionitis, may act in an additive or synergistic manner with postnatal insults, such as hyperoxia, ventilator-induced lung injury, or infection.^1 20 21^ Therefore, BPD is a condition with a multifactorial etiopathogenesis and the various factors that may favor or reduce BPD may be affected by sex differences. We will discuss the available evidence on these differences in the following paragraphs.

Since BPD is a condition that exclusively affects the developing lung, sexual dimorphism in the pattern of pulmonary development is often proposed as a potential explanation for sex differences in BPD rates. ^22–24^ A substantial body of evidence derived from preclinical and clinical studies indicates that female fetuses exhibit more advanced anatomical lung development and functional adaptation with regard to surfactant production than male fetuses of an identical GA.^22–27^ The most plausible explanation for these sex differences is the concomitant negative effect of androgens and the positive effect of estrogens and progesterone on structural lung development and surfactant synthesis.^23 24 28 29^ The relatively advanced lung development in preterm girls may account the lower rate of respiratory complications at birth and the lower need for mechanical ventilation, respiratory support, and surfactant when compared with preterm boys.^5 22 23^

Focusing on prenatal conditions, there is increasing attention to the sex-specific interface between the mother, fetus, and placenta in various pregnancy complications leading to sex-differences in neonatal outcomes.^30-35^ The two main pathophysiologic pathways, or endotypes, associated with extreme preterm birth are infection/inflammation and placental dysfunction.^36-38^ The first group includes chorioamnionitis, while the second group includes hypertensive disorders of pregnancy and the entity identified as fetal indication/intrauterine growth restriction.^30 38^ These two endotypes not only trigger preterm birth, but also may affect fetal lung development and may create a different susceptibility to postnatal pulmonary injury and therapies.^37–39^ Data from previous meta-analyses by our group suggest no association between chorioamnionitis and fetal sex.^35 40^ However, when chorioamnionitis was accompanied by funisitis, 1. the histologic equivalent of fetal inflammatory response syndrome 1. we observed an association with female fetal sex.^41^ With regard to the endotype of placental dysfunction, evidence shows that early-onset (< 32 weeks) hypertensive disorders of pregnancy are more frequently associated with female foetuses.^34 35^ Furthermore, the lesions indicative of maternal vascular malperfusion appear to be more pronounced in preeclamptic placentas derived from pregnancies with a female fetus compared to those with a male fetus.^30^ Interestingly, Chatterjee et al. showed that placentas from small for GA (SGA) males differentially expressed genes of immune response and inflammation while placentas from SGA females differentially expressed genes of organogenesis-related pathways such as angiogenesis, glycoproteins/extracellular matrix, cellular adhesion, and motility.^42^

At birth, the immature lung of the preterm infant is forced to continue its developmental trajectory without the protective effects of high levels of placental estrogen and progesterone. In addition, the lung is subjected to further damage from exposure to extrauterine life and eventual life-saving medical interventions. Preterm girls may be better protected against lung damage due to their higher antioxidant capacity compared to boys.^5 43^ Furthermore, it has been postulated that chromosomal sex may be associated with sexual dimorphism in the reparative response to postnatal pulmonary injury.^29^ Finally, infectious/inflammatory processes, such as sepsis or necrotizing enterocolitis, are more common in boys than in girls and can exacerbate lung damage.^5^ This may contribute to the observed sex differences in the development of BPD.

The only outcome for which evidence in favor of H_0_ (no sex differences) was found was mild BPD. However, it should be noted that the definition of mild BPD is based on the need for oxygen for 28 days after birth, but not at 36 weeks PMA. The 28-day criterion has been eliminated in the most recent definitions and classifications of BPD, which only consider respiratory status at 36 weeks PMA.^9 44^ Consequently, our classification of mild BPD is no longer used. The requirement for oxygen for 28 days postnatal may be more a consequence of prematurity than of lung damage, which may explain the absence of sex differences in this subgroup of BPD. It is noteworthy that our previous analysis of sex differences in the risk of developing retinopathy of prematurity also revealed that the male disadvantage was only apparent in the severe forms of the condition.^4^

The main limitation of our meta-analysis was the high heterogeneity. To investigate potential sources of heterogeneity, we performed a subgroup analysis based on geographic and sociodemographic settings and mean or median GA of the different cohorts included in the meta-analysis. The subgroup analysis suggested that the male disadvantage for BPD is less evident in countries with a lower SDI. Notably, in our previous meta-analysis on male disadvantage in severe ROP, we observed that this phenomenon was not present in certain regions of Asia and in the middle and middle-low SDI cohorts.^4^

With regard to GA, the results of the subgroup analysis indicated that both the effect size (expressed as RR or OR) and the strength of evidence (expressed as BF_10_) of the male disadvantage in BPD were lower in those cohorts with lower mean or median GA. These findings are consistent with those of Farstad et al.^45^ and Dassios et al.^46^, who employed a frequentist approach and reported that sex differences in BPD were not statistically significant (p > 0.05) in the lowest GA groups. Taken together, these data suggest that potential sex differences in lung development and the reparative response to postnatal lung injury are not of sufficient magnitude to result in sex differences in BPD in the most immature infants. Therefore, the male disadvantage or female advantage may not be detectable when the risk for BPD is extremely high, because immaturity outweighs any potential relative advantage based on sex. However, it is important to note that none of the GA subgroup analyses yielded conclusive evidence (BF_10_< 1/3) in favor of H_0_ (absence of sex differences). Consequently, our data in the lowest GA subgroups should be interpreted as absence of conclusive evidence and not as evidence of absence of sex differences.

BPD is a condition that is arbitrarily defined by its treatment rather than by its pathophysiology or clinical picture. This potentially has resulted in the lumping together of infants with diverse respiratory pathologies and subsequent long-term outcomes.^1 47^ There is a growing recognition of the clinical importance of pulmonary vascular disease and the impact of PH on the clinical course and outcomes of BPD.^3^ The so-called vascular phenotype of BPD is characterized by the presence of BPD-PH.^23^ A noteworthy finding of our study is the absence of evidence indicating sex differences in BPD-PH. Our findings are consistent with those of a recent retrospective, multicenter cohort study of 1156 infants with severe BPD.^48^ This study demonstrated the absence of sex-differences in terms of use of PH medication, need for tracheostomy, level of respiratory support at hospital discharge, or mortality. The authors speculate that sex may have less of an impact on modulating disease trajectory in infants with established severe BPD.^48^ Accordingly, the present data suggest that females are less likely to develop moderate to severe BPD, but may be equally as susceptible as males to develop the vascular phenotype of BPD. BPD-PH has been associated with the endotype of placental vascular dysfunction^19 49^ As discussed above, this endotype affects female fetuses more frequently than male fetuses.^20 34 35^ Furthermore, the placenta with vascular dysfunction appears to exhibit sexual dimorphism with respect to mediators of angiogenesis.^50 42^ This may result in an increased risk of altered pulmonary vascular development in female fetuses. Finally, it should be noted that there is a strong female disadvantage in the risk of developing adult pulmonary arterial hypertension.^51^ Yet, the severity and prognosis of the disease are worse in male than in female adults. Sex chromosomes play a key role in the sexual dimorphism of adult pulmonary arterial hypertension.^52 53 54^ BPD-PH might be the exception to the rule of male disadvantage in moderate-to-severe BPD due to the contribution of a similar sexual dimorphism of pulmonary vascular disease in extremely preterm infants.

As mentioned in the introduction, the survival rate of extremely preterm infants has improved in recent years, particularly in high-income countries. It has been proposed that the male disadvantage of prematurity is decreasing due to the greater impact of outcome improvements on boys compared to girls.^6^ However, this seems to be the case for mortality but not for other complications of prematurity, as we show here for BPD or have previously shown for retinopathy of prematurity.^4 5^ The temporal trend of BPD epidemiology may be influenced by a confluence of circumstances, including differences in the definition of BPD, increased survival among most immature infants who are inherently predisposed to BPD, differential use of oxygen and respiratory therapies, and the potential for targeted use of therapies to reduce the risk of BPD.^55^ Our data suggest that the confluence of these variables has not resulted in a discernible temporal shift in the male disadvantage of prematurity with regard to BPD.

## Conclusion

Although sex is an essential biological variable in determining the outcome of prematurity, it is rarely considered in the design of pre-clinical and clinical research, or in developing personalized medical strategies.^56 57^ Our study provides compelling evidence that males are at a significantly higher risk of developing moderate to severe BPD compared to females. However, this male disadvantage is much less apparent in the more immature infants and does not extend to the risk of developing pulmonary vascular disease associated with BPD. An important step in the process of individualizing the approach to BPD may be to consider the sex of the infant, as this information can be used to personalize care and potentially improve outcomes. Furthermore, as proposed by Lingappan et al., clinical trials should always consider sex as a biological variable in data collection, analysis, and reporting. Additionally, the interaction between intervention and sex should be evaluated by appropriate statistical methods.^56^ Finally, it is also essential to investigate the contribution of sex differences to pulmonary outcomes in former extremely preterm infants across the lifespan.^58 59^

## Data availability

All data relevant to the study are included in the article or uploaded as supplementary information.

## Data Availability

All data produced in the present work are contained in the manuscript

## Contributors

EV conceived and designed the study with input from the other authors. EvW-K and TMH executed the literature search, screened and reviewed the search results, abstracted the data, and assessed the risk of bias. EV and KVM checked data extraction for accuracy and completeness. FB and EV designed and conducted the analysis. All authors contributed to the interpretation of analysis. EV, EvW-K and TMH made the figures and tables. EV drafted the manuscript with input from the other authors. All authors reviewed the manuscript and provided important intellectual content. EV and EvW-K take responsibility for the article as a whole.

## Ethics statement

As this systematic review and meta-analysis did not involve animal subjects or personally identifiable information on human subjects, ethics review board approval and patient consent were not required.

