## Supplementary material for "Sex Differences in the Risk of Bronchopulmonary Dysplasia and Pulmonary Hypertension: A Bayesian Meta-Analysis"

**1. Methods**

**1.1. Search strategy**

**PubMed**

((Sex[MESH] OR Sex Characteristics[MESH] OR Sex Distribution[MESH] OR "Sex Characteristic*"[tiab] OR "Gender Difference*"[tiab] OR "Sex Dimorphism*"[tiab] OR "Gender Characteristic*"[tiab] OR "Sex Difference*"[tiab] OR "Sex Distribution*"[tiab] OR "Gender Distribution*"[tiab] OR "Male Disadvantage"[tiab] OR "Female Advantage"[tiab] OR risk [tiab]) AND (Bronchopulmonary dysplasia[MESH] OR "BPD"[tiab] OR "chronic lung disease"[tiab] OR "CLD"[tiab] OR "pulmonary hypertension"[tiab]))

**Embase**

((exp Sex Characteristics/ OR exp Sex Distribution / OR exp Sex Factors/) OR ("male disadvantage" or "female advantage" or gender or sex or "gender difference" or "gender differences" or "sex difference" or "sex differences" or "gender differential" or "gender differentials" or "sex differential" or "sex differentials" or "sexual dimorphism" or "sexually dimorphic") ) AND (bronchopulmonary dysplasia))

No language limits were set. Narrative reviews, systematic reviews, case reports, letters, editorials, and commentaries were excluded, but read to identify potential additional studies. Additional strategies to identify studies included manual review of reference lists from key articles that fulfilled our eligibility criteria, use of “related articles” feature in PubMed, and use of the “cited by” tool in Web of Science and Google scholar. Two reviewers independently screened the results of the searches, and included studies according to the inclusion criteria using EndNote (RRID:SCR_014001), using the methodology described by Bramer et al.^1^

**1.2. Supplementary information on methods**

*Study selection*

We selected cohort studies in which sex was the independent variable and the outcome (BPD or BPD-PH) the dependent variable as well as studies in which the outcome was the independent variable and sex the dependent variable. Studies that exclusively included late preterm infants (GA ≥34 weeks) or that combined preterm and term infants were excluded. The absence of a clear definition of BPD was also an exclusion criterion. Due to the high number of included studies, no additional efforts were made to clarify the definitions or other data with the authors. Abstracts and unpublished studies were also excluded.

*Data extraction*

Data extracted from each study included citation information, language of publication, location where research was conducted, sociodemographic index (SDI), time period of the study, study objectives, study design, inclusion/exclusion criteria, definition criteria for BPD, and BPD-PH, patient characteristics, and results (including raw numbers or summary statistics when raw numbers were not available). The SDI is a composite measure of developmental status as it is associated with health outcomes, calculated as the geometric mean of the following three indicators: total rate of fertility, log income per capita, and mean years of education among those 15 years or older. SDI values are scaled from 0 (highest fertility, lowest income, and lowest education) to 1 (highest income, highest education, and lowest fertility).^2^ For comparisons across SDI quintiles, each country was assigned to a single quintile according to its SDI in 2019.

*Assessment of risk of bias*

Methodological quality was assessed using the Newcastle‒Ottawa scale (NOS) for cohort studies.^3^ This scale assigns a maximum of 9 points (4 for selection, 2 for comparability, and 3 for outcome). NOS scores ≥ 7 were considered high-quality studies (low risk of bias), and scores of 5 to 6 denoted moderate quality (moderate risk of bias) ^2^. Risk of bias was additionally assessed using the Quality in Prognosis Studies (QUIPS) tool, with the modifications proposed by Stallings et al. for the study of sex as a prognostic factor.^4^ The tool consists of several questions within six different domains (study participation, study attrition, sex assessment, other prognostic factor adjustment, outcome measurement and analysis/ reporting), with each domain rated on a four-point scale.^4^

*Categories of evidence*

The BF_10_ is the ratio of the probability of the data under H_1_ over the probability of the data under H_0_. The BF_10_ was interpreted using the evidence categories suggested by Lee & Wagenmakers.^5^ The evidence in favor of H_1_ (BF_10_ > 1) was categorized as weak/inconclusive (1< BF_10_ < 3), moderate (3< BF_10_ < 10), strong (10< BF_10_ < 30), very strong (30< BF_10_< 100), and extreme (BF_10_ >100). The evidence in favor of H_0_ (BF_10_ < 1) was categorized as weak/inconclusive (1/3< BF_10_ < 1), moderate (1/10< BF_10_ < 1/3), strong (1/30< BF_10_ < 1/10), very strong (1/100< BF_10_< 1/30), and extreme (BF_10_ <1/100).The BF_rf_ is the ratio of the probability of the data under the random effects model over the probability of the data under the fixed effect model. The categories of strength of the evidence in favor of the random effects (BF_rf_ > 1) or the fixed effect (BF_rf_< 1) were similar to those described above for BF_10_.

*Robust Bayesian meta-analysis (RoBMA)*

We used RoBMA to assess the robustness of the results to the potential presence of publication bias.^6^ RoBMA extends the Bayesian model-averaged meta-analysis by the two major publication bias adjustment techniques: selection models (adjusting for the publication bias operating on *p*-values) and precision-effect test and precision-effect estimate with standard errors (PET-PEESE, adjusting for the relationship between effect sizes and standard errors).^7^ The resulting RoBMA ensemble contains 36 models composed of the following assumptions about the presence vs. absence of the effect (2) x presence vs. absence of between-study heterogeneity (2) x presence vs. absence of publication bias adjustment models (6 selection models, PET, PEESE, and no bias). We used RoBMA with the same prior distributions for the effect and heterogeneity as in BMA and the default prior distributions for the publication bias adjustment part. Publication bias was expressed as BF_bias_ using the same categories for evidence previously described for BF_10_ and BF_rf_.

*Meta-regression*

We used meta-regression analyses to test whether there was a significant relationship between the median year of birth of the the cohort and effect size, as indicated by a Z-value and an associated p-value. Meta-regression coefficient indicates the change in the log of the RR of the association between BPD and the corresponding exposure for a unit change in the predictor covariate. Subgroups were compared using meta-regression for categorical covariates. For both categorical and continuous covariates, the R^2^ analog, defined as the total between-study variance explained by the moderator, was calculated based on the meta-regression matrix. ^8^

**2. Results**

**2.1. Supplementary Figures**

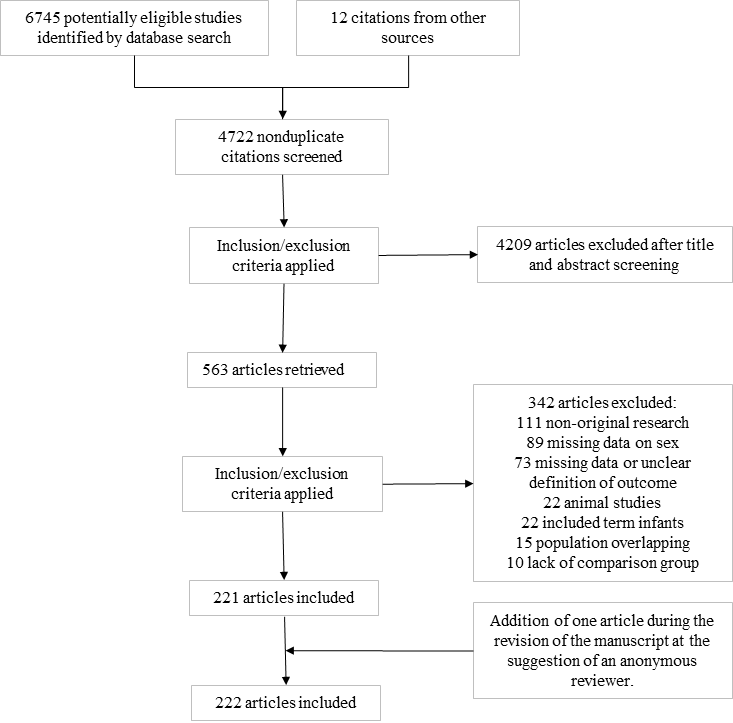

**Supplementary Figure 1.** Flow diagram of the systematic search.

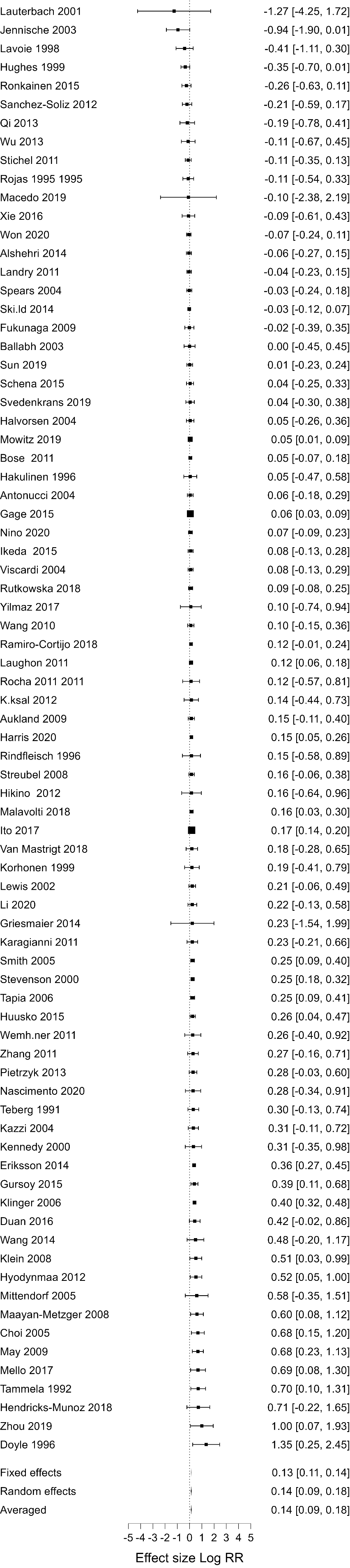

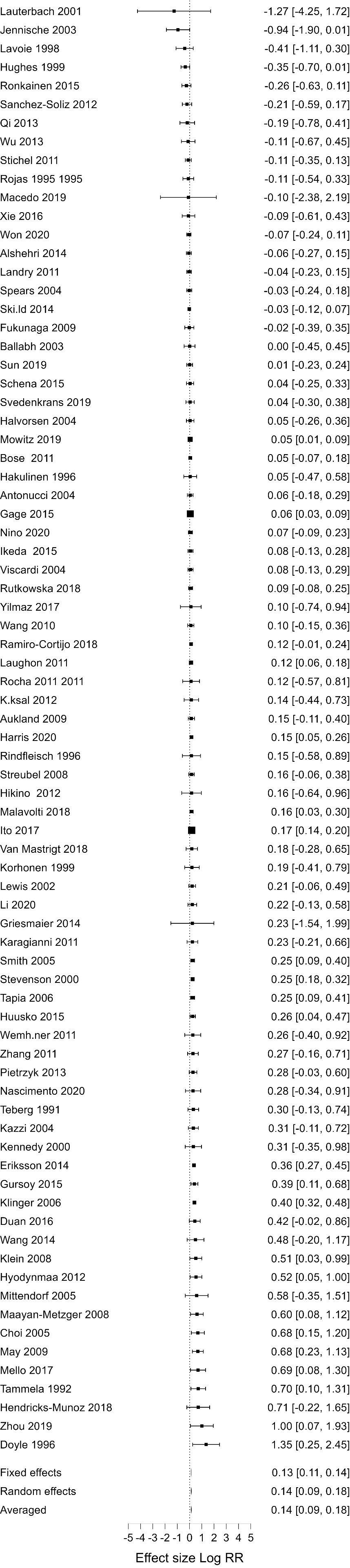

**Supplementary Figure 2.** Forest plot of Bayesian model averaged (BMA) meta-analysis on the association between infant sex and bronchopulmonary dysplasia defined as oxygen requirement during the first 28 days of life or at postnatal day 28 (BPD28). Log RR> 0 indicates higher risk in males.

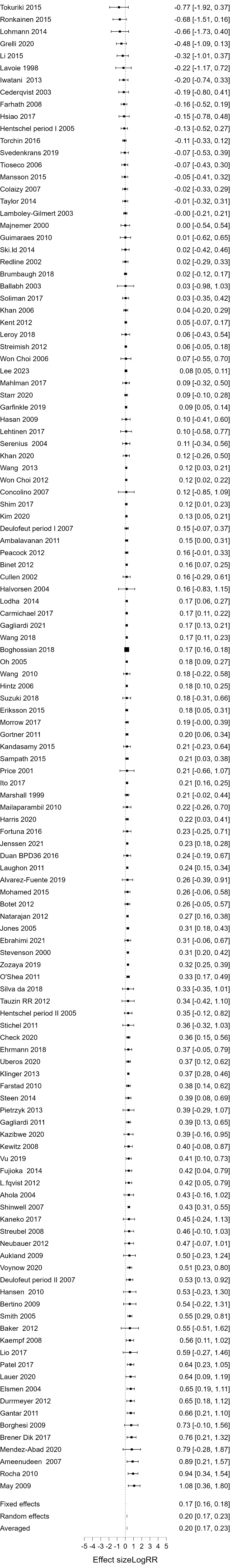

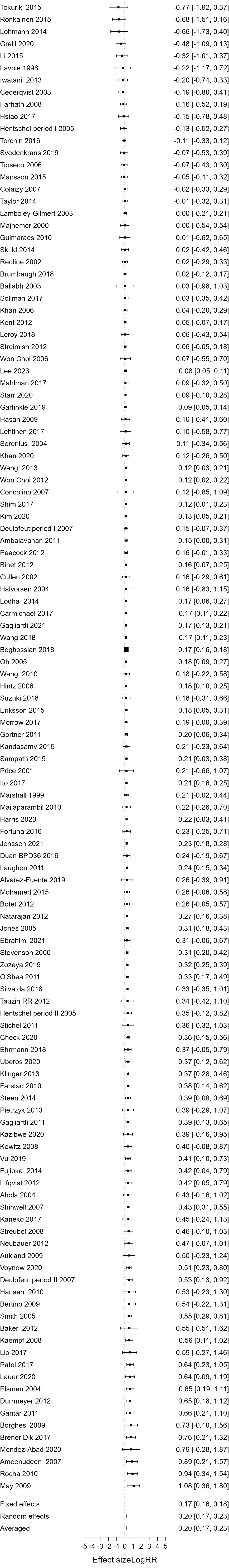

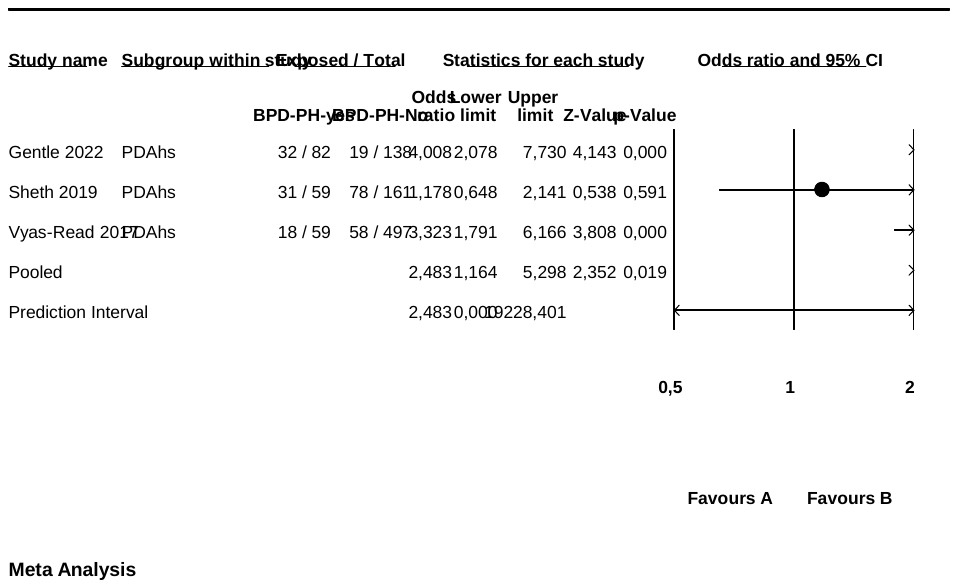

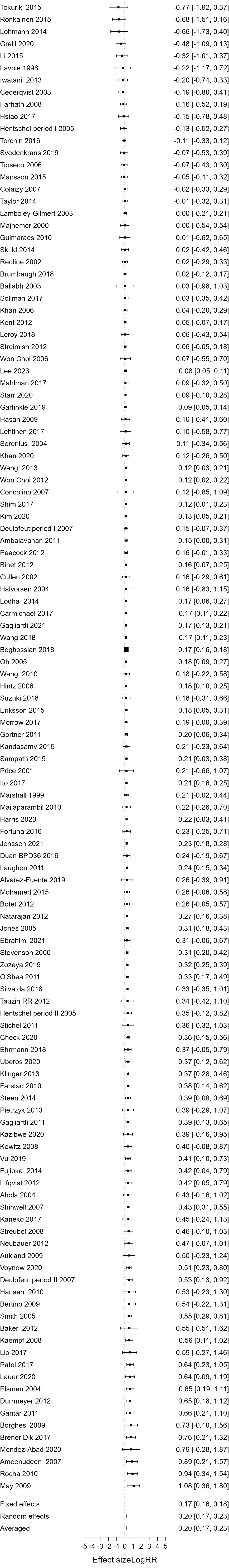

**Supplementary Figure 3.** Forest plot of Bayesian model averaged (BMA) meta-analysis on the association between infant sex and bronchopulmonary dysplasia defined as oxygen or respiratory support requirement at the post-menstrual age of 36 weeks (BPD36). Log RR> 0 indicates higher risk in males.

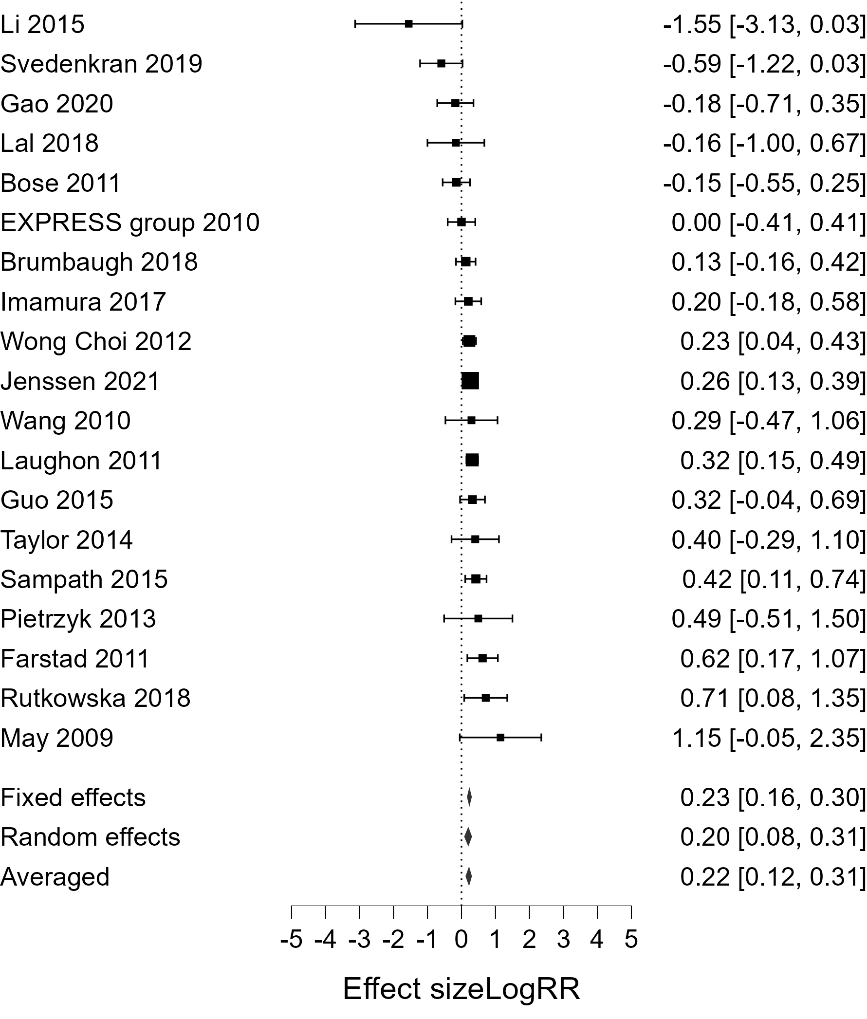

**Supplementary Figure 4.** Forest plot of Bayesian meta-analysis on the association between infant sex and severe bronchopulmonary dysplasia. Log RR> 0 indicates higher risk in males.

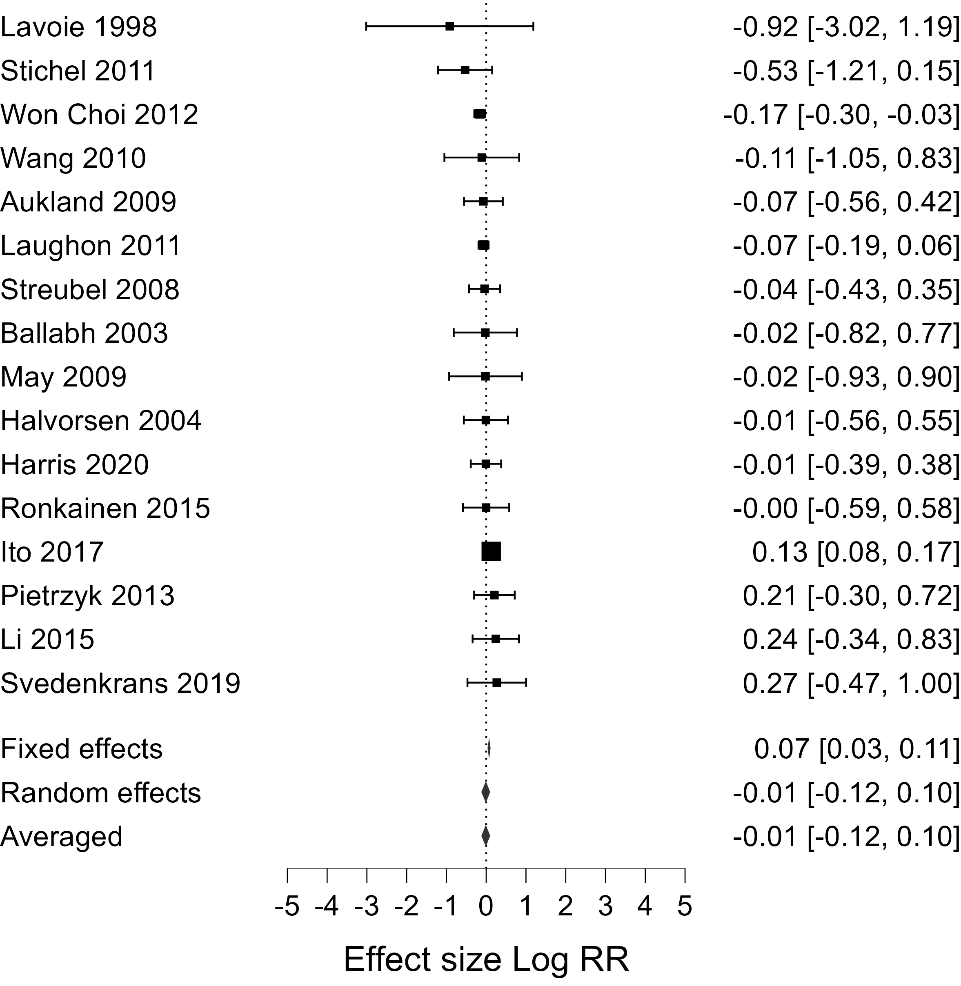

**Supplementary Figure 5.** Forest plot of Bayesian model averaged (BMA) meta-analysis on the association between infant sex and mild bronchopulmonary dysplasia. Log RR> 0 indicates higher risk in males.

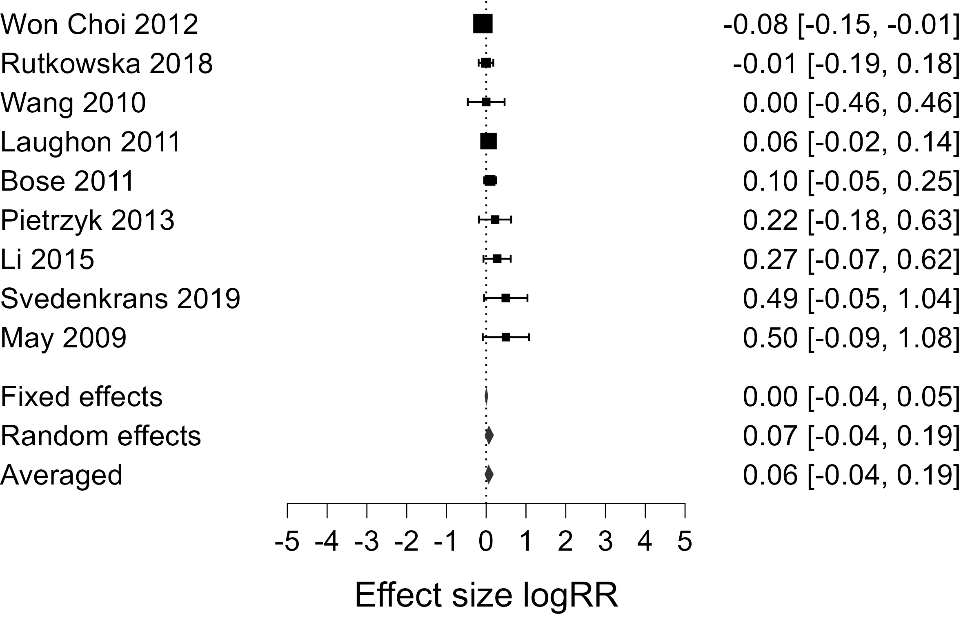

**Supplementary Figure 6.** Forest plot of Bayesian model averaged (BMA) meta-analysis on the association between infant sex and mild/moderate bronchopulmonary dysplasia. Log RR> 0 indicates higher risk in males.

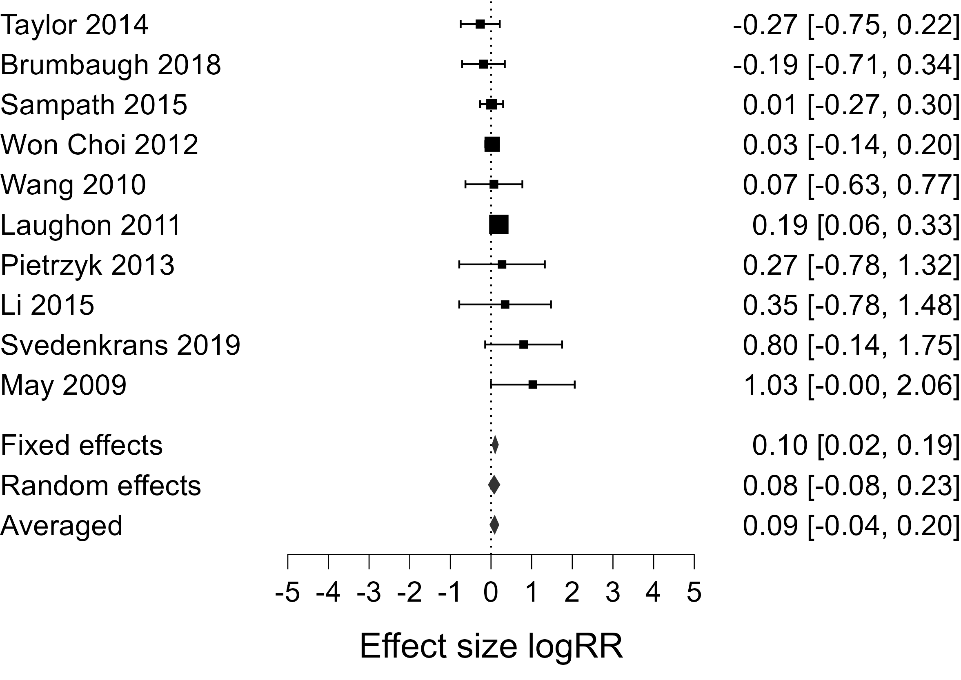

**Supplementary Figure 7.** Forest plot of Bayesian model averaged (BMA) meta-analysis on the association between infant sex and moderate bronchopulmonary dysplasia. Log RR> 0 indicates higher risk in males.

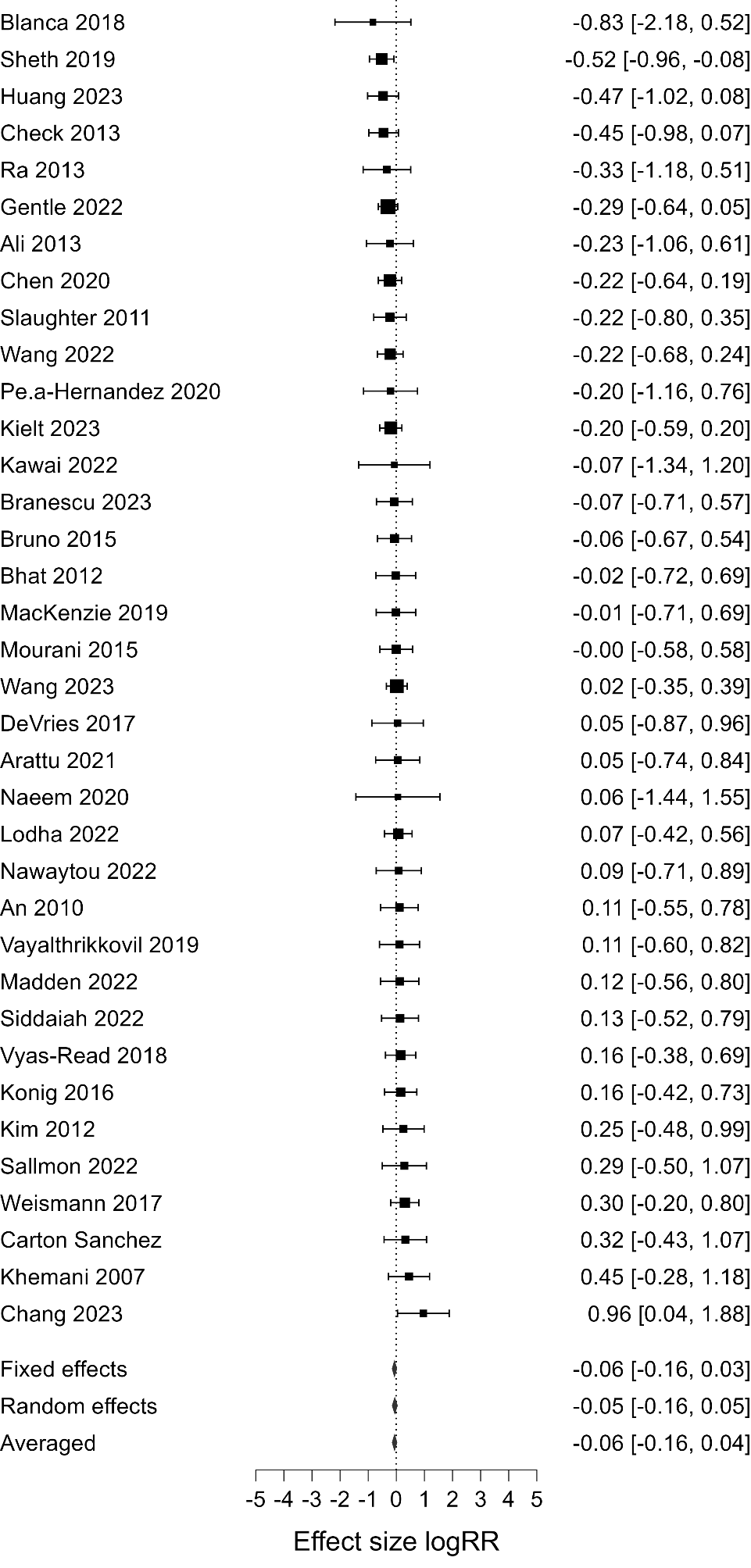

**Supplementary Figure 8.** Forest plot of Bayesian model averaged (BMA) meta-analysis on the association between infant sex and bronchopulmonary dysplasia-associated pulmonary hypertension. Log RR> 0 indicates higher risk in males.

**2.2. Supplementary Tables**

**Supplementary Table 1.** Characteristics of the included studies and risk of bias assessment

| First author, Year | Country | Median Year | Prospective? | Independent variable | Total infants (centers) | Mean GA of cohort | Outcomes | NOS |
| --- | --- | --- | --- | --- | --- | --- | --- | --- |
| Ahola 2004 ^9^ | Finland | NA | Yes | BPD | 83 (10) | 26.7 | BPD36 | 7 |
| Ali 2013 ^10^ | Denmark | 2006 | No | BPD | 74 (1) | 26.6 | BPD-PH | 9 |
| Alshehri 2014^11^ | Saudi Arabia | 2007 | No | BPD | 942 (1) | 29.3 | BPD28 | 7 |
| Alvarez-Fuente 2019 ^12^ | Spain | 2015 | Yes | BPD | 47 (5) | 26.0 | BPD36 | 8 |
| Ambalavanan 2011 ^13^ | USA | 2004 | No | BPD | 5630 (17) | 26.9 | BPD36 | 8 |
| Ameenudeen 2007^14^ | Malaysia | 2004 | Yes | BPD | 244 (1) | 29.8 | BPD36 | 9 |
| An 2010 ^15^ | Korea | 2006 | No | BPD | 116 (1) | 26.3 | BPD-PH | 8 |
| Antonucci 2004 ^16^ | Italy | 1996 | No | BPD | 277 (1) | 30.1 | BPD28 | 9 |
| Arattu 2021^17^ | UK | 2019 | No | BPD | 182 (1) | 26.2 | BPD-PH | 8 |
| Aukland 2009 ^18^ | Norway | 1987 | Yes | BPD | 74 (1) | 26.9 | BPD28 & BPD36  Mild BPD | 8 |
| Baker 2012^19^ | USA | 2010 | Yes | BPD | 62 (1) | 31.6 | BPD36 | 7 |
| Ballabh 2003 ^20^ | USA | 2000 | Yes | BPD | 39 (1) | 26.7 | BPD28 & BPD36  Mild BPD | 7 |
| Bertino 2009 ^21^ | Italy | 1997 | Yes | sex | 262 (1) | 30.4 | BPD36 | 9 |
| Bhat 2012 ^22^ | USA | 2010 | Yes | BPD | 145 (1) | 25.7 | BPD-PH | 9 |
| Binet 2012 ^23^ | Canada | 2003 | No | sex | 2744 (46) | 25.8 | BPD36 | 7 |
| Blanca 2018 ^24^ | The Netherlands | 2015 | Yes | BPD | 69 (1) | 25.7 | BPD-PH | 7 |
| Boghossian 2018 ^25^ | USA | 2011 | No | sex | 161601 (766) | 27.5 | BPD36 | 7 |
| Borghesi 2009 ^26^ | Italy | 2008 | Yes | BPD | 98 (1) | 29.5 | BPD36 | 9 |
| Bose 2011^27^ | USA | 2003 | Yes | BPD | 932 (14) | 26.0 | BPD28  Mild/Mod BPD  Sev BPD | 6 |
| Botet 2012 ^28^ | Spain | 2003 | No | BPD | 415 (1) | 27.0 | BPD36 | 9 |
| Branescu 2023 ^29^ | UK | 2016 | No | BPD | 268 (1) | 26.6 | BPD-PH |  |
| Brener Dik 2017^30^ | Argentina | 2012 | No | BPD | 203 (1) | 28.8 | BPD36 | 9 |
| Brumbaugh 2018 ^31^ | USA | 2011 | No | BPD | 151 (1) | 26.6 | BPD36  Mod BPD  Sev BPD | 9 |
| Bruno 2015 ^32^ | USA | 2009 | No | BPD | 303 (1) | 26.6 | BPD-PH | 9 |
| Carmichael 2017 ^33^ | USA | 2009 | No | BPD | 10182 (136) | 27.5 | BPD36 | 8 |
| Carton-Sanchez 2016 ^34^ | Spain | 2011 | Yes | BPD | 84 (1) | 27.0 | BPD-PH | 9 |
| Cederqvist 2003^35^ | Finland | 1995 | Yes | BPD | 32 (1) | 27.3 | BPD36 | 6 |
| Chang 2023 ^36^ | Taiwan | 2020 | No | BPD | 139 (1) | 27.3 | BPD-PH |  |
| Check 2013 ^37^ | USA | 2007 | No | BPD | 138 (1) | 26.1 | BPD-PH | 7 |
| Check 2020 ^38^ | USA | NA | Yes | BPD | 1149 (2) | 27.4 | BPD36 | 9 |
| Chen 2020 ^39^ | USA | NA | Yes | BPD | 188 (1) | 26.7 | BPD-PH |  |
| Choi 2005^40^ | Korea | 1998 | Yes | BPD | 115 (1) | 29.9 | BPD28 | 9 |
| Colaizy 2007^41^ | USA | 2000 | Yes | BPD | 121 (1) | 26.9 | BPD36 | 7 |
| Concolino 2007 ^42^ | Italy | 2006 | Yes | BPD | 33 (1) | 27.7 | BPD36 | 6 |
| Cullen 2002 ^43^ | USA | 1998 | Yes | BPD | 132 (1) | 27.0 | BPD36 | 7 |
| Deulofeut 2007 ^44^ | USA | 2003 | No | sex | 497 (2) | 27.1 | BPD36 | 9 |
| DeVries 2017^45^ | USA | 2008 | No | BPD | 577 (1) | 26.6 | BPD-PH | 8 |
| Doyle 1996 ^46^ | Australia | 1981 | Yes | BPD | 154 (1) | 29.4 | BPD28 | 9 |
| Duan 2016a ^47^ | China | 2015 | Yes | BPD | 243 (1) | 30.0 | BPD28 | 9 |
| Duan 2016b ^48^ | China | 2015 | Yes | BPD | 147 (1) | 29.5 | BPD36 | 9 |
| Durrmeyer 2012^49^ | France | 2004 | No | BPD | 351 (1) | 26.5 | BPD36 | 7 |
| Ebrahimi 2021 ^50^ | The Netherlands | 2012 | No | BPD | 209 (1) | 27.3 | BPD36 | 8 |
| Ehrmann 2018 ^51^ | USA | 2009 | No | BPD | 93 (NA) | 27.8 | BPD36 | 9 |
| Elsmen 2004 ^52^ | Sweden | 1997 | No | sex | 236 (1) | 26.3 | BPD36 | 7 |
| Eriksson 2014^53^ | Sweden | 1999 | No | BPD | 106339 (NA) | <37 | BPD28 | 7 |
| Eriksson 2015 ^54^ | Sweden | 2008 | No | BPD | 2255 (NA) | <37 | BPD36 | 9 |
| EXPRESS group 2010^55^ | Sweden | 2006 | Yes | BPD | 497 (NA) | 25.0 | Sev BPD | 9 |
| Farhath 2008 ^56^ | USA | 2005 | Yes | BPD | 43 (1) | 26.0 | BPD36 | 7 |
| Farstad 2011 ^57^ | Norway |  |  |  |  |  |  |  |
| Fortuna 2016 ^58^ | Italy | 2001 | Yes | BPD | 48 (NA) | 25.9 | BPD36 | 7 |
| Fujioka 2014 ^59^ | Japan | 2007 | No | BPD | 97 (1) | 27.9 | BPD36 | 7 |
| Fukunaga 2009 ^60^ | Japan | 2005 | No | BPD | 29 (1) | 27.8 | BPD28 | 7 |
| Gage 2015 ^61^ | USA | 2009 | No | BPD | 21944 (NA) | <37 | BPD28 | 8 |
| Gagliardi 2011 ^62^ | Italy | 2001 | Yes | BPD | 1260 (14) | 29.0 | BPD36 | 9 |
| Gagliardi 2021 ^63^ | Multi | 2011 | No | sex | 17982 (NA) | 26.9 | BPD36 | 8 |
| Gantar 2011 ^64^ | Slovenia | 2001 | Yes | BPD | 115 (1) | 27.3 | BPD36 | 7 |
| Gao 2020 ^65^ | China | 2016 | No | BPD | 155 (1) | 28.6 | Sev BPD | 7 |
| Garfinkle 2019 ^66^ | Canada | 2008 | No | sex | 15153 (NA) | 26.0 | BPD36 | 9 |
| Gentle 2022 ^67^ | USA | 2019 | Yes | BPD | 220 (1) | 25.6 | BPD-PH | 9 |
| Gortner 2011^68^ | Europe | 2003 | Yes | BPD | 4185 (NA) | 30.0 | BPD36 | 8 |
| Grelli 2020 ^69^ | USA | 2014 | No | BPD | 87 (1) | 28.1 | BPD36 | 9 |
| Griesmaier 2014 ^70^ | Austria | 2010 | Yes | sex | 156 (1) | 30.5 | BPD28 | 7 |
| Guimaraes 2010^71^ | Portugal | 2005 | Yes | BPD | 256 (5) | 28.6 | BPD36 | 7 |
| Guo 2015^72^ | Taiwan | 2009 | No | BPD | 75 (1) | 27.6 | SEVERE | 7 |
| Gursoy 2015 ^73^ | Turkey | 2008 | No | BPD | 652 (1) | 29.4 | BPD28 | 9 |
| Hakulinen 1996 ^74^ | Finland | 1982 | No | BPD | 31 (1) | 28.0 | BPD28 | 7 |
| Halvorsen 2004 ^75^ | Norway | 1984 | No | BPD | 46 (1) | 27.3 | BPD28 & BPD36  Mild BPD | 7 |
| Hansen 2010^76^ | USA | 2007 | Yes | BPD | 107 (1) | 29.0 | BPD36 | 9 |
| Harris 2020 ^77^ | UK | NA | No | sex | 319 (1) | 26.8 | BPD28 & BPD36  Mild BPD | 9 |
| Hasan 2009 ^78^ | USA | 2006 | Yes | BPD | 65 (NA) | 27.0 | BPD36 | 7 |
| Hendricks-Munoz 2018 ^79^ | USA | NA | Yes | BPD | 25 (1) | 26.1 | BPD28 | 6 |
| Hentschel 2005 ^80^ | Switzerland | 1998 | Yes | BPD | 1274 (9) | 29.5 | BPD36 |  |
| Hikino 2012^81^ | Japan | NA | Yes | BPD | 26 (1) | 29.0 | BPD28 | 7 |
| Hintz 2006 ^82^ | USA | 1999 | No | sex | 2552 (NA) | 25.5 | BPD36 | 9 |
| Hsiao 2017 ^83^ | Taiwan | NA | Yes | BPD | 80 (1) | 27.9 | BPD36 | 7 |
| Huang 2023 ^84^ | Taiwan | 2019 | No | BPD | 91 (1) | 25.7 | BPD-PH |  |
| Hughes 1999 ^85^ | USA | 1980 | Yes | BPD | 407 (NA) | 30.1 | BPD28 | 8 |
| Huusko 2015 ^86^ | Canada | 2000 | Yes | BPD | 841 (8) | 28.4 | BPD28 | 9 |
| Hyödynmaa 2012 ^87^ | Finland | NA | Yes | BPD | 82 (1) | 29.4 | BPD28 | 6 |
| Ikeda 2015 ^88^ | Japan | 2007 | Yes | BPD | 294 (1) | 27.7 | BPD28 | 7 |
| Imamura 2017 ^89^ | Japan | 2010 | No | BPD | 169 (1) | 26.0 | Sev BPD | 9 |
| Ito 2017 ^90^ | Japan | 2008 | No | sex | 38023 (NA) | 28.2 | BPD28 & BPD36  Mild BPD | 7 |
| Iwatani 2013^91^ | Japan | 2009 | No | BPD | 51 (1) | 25.6 | BPD36 | 7 |
| Jennische 2003 ^92^ | Sweden | 1988 | No | sex | 64 (1) | 28.9 | BPD28 | 9 |
| Jensen 2021^93^ | USA | 2018 | No | BPD | 24896 (715) | 27.1 | BPD36  SevBPD | 9 |
| Jones 2005 ^94^ | Canada | 1997 | Yes | sex | 3398 (17) | 28.0 | BPD36 | 9 |
| Kaempf 2008 ^95^ | USA | NA | Yes | BPD | 220 (8) | 27.8 | BPD36 | 7 |
| Kandasamy 2015^96^ | USA | 2007 | Yes | BPD | 152 (1) | 25.2 | BPD36 | 7 |
| Kaneko 2017 ^97^ | Japan | 2011 | No | BPD | 31 (1) | 26.4 | BPD36 | 8 |
| Karagianni 2011^98^ | Greece | 2007 | Yes | BPD | 219 (1) | 29.1 | BPD28 | 7 |
| Kawai 2022 ^99^ | Japan | 2015 | No | BPD | 131 (1) | 26.6 | BPD-PH | 9 |
| Kazibwe 2020 ^100^ | USA | 2012 | No | BPD | 165 (1) | 29.0 | BPD36 | 7 |
| Kazzi 2004^101^ | USA | 2002 | Yes | BPD | 120 (1) | 28.2 | BPD28 | 7 |
| Kennedy 2000 ^102^ | Australia | 1982 | No | BPD | 102 (1) | 29.6 | BPD28 | 9 |
| Kent 2012 ^103^ | Australia | 2001 | No | sex | 2549 (10) | 26.3 | BPD36 | 9 |
| Kewitz 2008 ^104^ | Germany | 1999 | Yes | BPD | 115 (1) | 26.2 | BPD36 | 8 |
| Khan 2006^105^ | USA | 1997 | No | BPD | 306 (2) | 26.9 | BPD36 | 7 |
| Khan 2020 ^106^ | USA | NA | Yes | BPD | 68 (1) | 26.7 | BPD36 | 9 |
| Khemani 2007 ^107^ | USA | 2002 | No | BPD | 42 (3) | 26.5 | BPD-PH | 7 |
| Kielt 2023 ^108^ | USA | 2014 | No | BPD | 223 (1) | 25.0 | BPD-PH | 8 |
| Kim 2012 ^109^ | Korea | 2007 | No | BPD | 98 (1) | 26.8 | BPD-PH | 8 |
| Kim 2020 ^110^ | Korea | 2015 | No | BPD | 3039 (NA) | 26.4 | BPD36 | 8 |
| Klein 2008 ^111^ | Argentina | 2004 | Yes | sex | 119 (2) | 29.8 | BPD28 | 9 |
| Klinger 2006 ^112^ | Israel | 1999 | Yes | BPD | 10134 (28) | 27.5 | BPD28 | 9 |
| Klinger 2013^113^ | Israel | 2005 | Yes | BPD | 12139 (28) | 28.5 | BPD36 | 9 |
| Köksal 2012 ^114^ | Turkey | 2010 | Yes | BPD | 102 (1) | 28.6 | BPD28 | 7 |
| Konig 2016 ^115^ | Australia | NA | No | BPD | 83 (1) | 26.1 | BPD-PH | 7 |
| Korhonen 1999 ^116^ | Finland | 1992 | Yes | BPD | 143 (1) | 28.5 | BPD28 | 8 |
| Lal 2018 ^117^ | USA | 2002 | Yes | BPD | 30 (1) | 24.5 | SEVERE | 7 |
| Lamboley-Gilmert 2003^118^ | France | 1995 | Yes | BPD | 667 (33) | 29 | BPD36 | 7 |
| Landry 2011 ^118^ | Canada | 1986 | No | BPD | 1192 (1) | 31.4 | BPD28 | 9 |
| Lauer 2020 ^120^ | Germany | 2016 | No | BPD | 102 (1) | 26.4 | BPD36 | 9 |
| Laughon 2011 ^121^ | USA | 2002 | Yes | BPD | 3629 (17) | 27.0 | BPD28 & BPD36  Mild BPD  Mild/Mod BPD  Mod BPD  Sev BPD | 9 |
| Lauterbach 2001 ^122^ | USA | 1990 | No | sex | 51 (1) | 32.3 | BPD28 | 6 |
| Lavoie 1998 ^123^ | Canada | 1988 | No | sex | 22 (1) | 27.0 | BPD28 & BPD36  Mild BPD | 6 |
| Lehtinen 2017 ^124^ | Finland | 2001 | Yes | BPD | 53 (1) | 29.0 | BPD36 | 9 |
| Lee 2023 ^125^ | UK | 2016 | No | sex | 9998 (NA) | 26.1 | BPD36 | 8 |
| Leroy 2018^126^ | Canada | NA | Yes | BPD | 62 (1) | 26.9 | BPD36 | 7 |
| Lewis 2002 ^127^ | USA | NA | Yes | BPD | 160 (3) | 28.3 | BPD28 | 7 |
| Li 2015^128^ | China | 2013 | No | BPD | 47 (NA) | 32.0 | BPD36  Mild BPD  Mild/Mod BPD  Mod BPD  Sev BPD | 7 |
| Li 2020 ^129^ | China | 2016 | No | BPD | 213 (1) | 30.2 | BPD28 | 7 |
| Lio 2017 ^130^ | Italy | 2011 | Yes | BPD | 154 (1) | 27.1 | BPD36 | 8 |
| Lodha 2014^131^ | Canada | 2001 | Yes | BPD | 1030 (1) | 28.5 | BPD36 | 6 |
| Lodha 2022 ^132^ | Canada | 2018 | No | BPD | 254 (1) | 25.8 | BPD-PH | 8 |
| Löfqvist 2012 ^133^ | Sweden | 2003 | No | BPD | 108 (2) | 27.2 | BPD36 | 9 |
| Lohmann 2014^134^ | USA | NA | Yes | BPD | 22 (1) | 27.7 | BPD36 | 7 |
| Maayan-Metzger 2008 ^135^ | Israel | 1997 | Yes | BPD | 325 (1) | 29.0 | BPD28 | 8 |
| Macedo 2019 ^136^ | Portugal | 2014 | Yes | sex | 32 (1) | 29.8 | BPD28 | 8 |
| MacKenzie 2019 ^137^ | Canada | 2014 | No | BPD | 92 (1) | 25.9 | BPD-PH | 8 |
| Madden 2022 ^138^ | USA | 2018 | No | BPD | 64 (1) | 25.9 | BPD-PH | 8 |
| Mahlman 2017^139^ | Finland | NA | Yes | BPD | 174 (5) | 27.4 | BPD36 | 7 |
| Mailaparambil 2010^140^ | Germany | 2002 | Yes | BPD | 155 (1) | 25.7 | BPD36 | 7 |
| Majnemer 2000 ^141^ | Canada | 1986 | Yes | BPD | 54 (1) | 29.5 | BPD36 | 6 |
| Malavolti 2018 ^142^ | Switzerland | 2007 | No | BPD | 610 (1) | 27.9 | BPD28 | 9 |
| Mansson 2015 ^143^ | Sweden | 2006 | Yes | sex | 398 (NA) | 25.5 | BPD36 | 9 |
| Marshall 1999 ^144^ | USA | 1994 | Yes | BPD | 865 (11) | 28.9 | BPD36 | 9 |
| May 2009 ^145^ | UK | NA | Yes | BPD | 80 (1) | 27.9 | Mild BPD  Mild/Mod BPD  Mod BPD  Sev BPD | 7 |
| Mello 2017 ^146^ | Brazil | 2006 | Yes | BPD | 112 (1) | 29.5 | BPD28 | 8 |
| Méndez-Abad 2020 ^147^ | Spain | 2016 | Yes | BPD | 101 (1) | 29.0 | BPD36 | 7 |
| Mittendorf 2005^148^ | USA | 1996 | Yes | BPD | 141 (1) | 29.9 | BPD28 | 7 |
| Mohamed 2015 ^149^ | Canada | 2011 | Yes | BPD | 99 (1) | 26.0 | BPD36 | 8 |
| Morrow 2017^150^ | USA | 2011 | Yes | BPD | 587 (5) | 26.6 | BPD36 | 9 |
| Mourani 2015 ^151^ | USA | 2009 | Yes | BPD | 274 (2) | 26.7 | BPD-PH | 8 |
| Mowitz 2019 ^152^ | USA | 2012 | No | BPD | 12017 (500) | 26.5 | BPD28 | 8 |
| Naeem 2020 ^153^ | USA | 2017 | Yes | BPD | 36 (1) | 26.0 | BPD-PH | 8 |
| Nascimento 2020 ^154^ | Brazil | 2016 | Yes | BPD | 40 (1) | 31.5 | BPD28 | 8 |
| Natarajan 2012 ^155^ | USA | 2007 | Yes | BPD | 1159 (NA) | 25.7 | BPD36 | 9 |
| Nawaytou 2022 ^156^ | USA | 2018 | Yes | BPD | 256 (1) | 26.2 | BPD-PH | 8 |
| Neubauer 2012 ^157^ | Austria | 2006 | Yes | sex | 408 (1) | 29.3 | BPD36 | 7 |
| Nino 2020 ^158^ | USA | NA | No | BPD | 188 (1) | 27.0 | BPD28 | 7 |
| O’Shea 2011 ^159^ | Australia | 1998 | Yes | BPD | 751 (3) | 26.6 | BPD36 | 9 |
| Oh 2005 ^160^ | USA | 2000 | No | BPD | 1382 (NA) | 25.8 | BPD36 | 9 |
| Patel 2017 ^161^ | USA | 2010 | Yes | BPD | 254 (1) | 27.9 | BPD36 | 9 |
| Peacock 2012 ^162^ | UK | 2000 | Yes | sex | 797 (NA) | 26.5 | BPD36 | 7 |
| Peña-Hernandez 2020 ^163^ | USA | 2000 | No | BPD | 143 (NA) | 31.3 | BPD-PH |  |
| Pietrzyk 2013 ^164^ | Poland | 2009 | Yes | BPD | 111 (1) | 27.8 | BPD28 & BPD36  Mild BPD  Mild/Mod BPD  Mod BPD  Sev BPD | 9 |
| Price 2001^165^ | USA | 1995 | Yes | BPD | 29 (1) | 28.3 | BPD36 | 9 |
| Qi 2013 ^166^ | China | 2011 | Yes | BPD | 60 (1) | 29.5 | BPD28 | 7 |
| Ra 2013 ^167^ | Korea | 2009 | No | BPD | 85 (1) | 28.0 | BPD-PH | 7 |
| Ramiro-Cortijo 2018 ^168^ | Spain | 2010 | No | sex | 1390 (1) | 28.2 | BPD28 | 8 |
| Redline 2002 ^169^ | USA | 1996 | No | BPD | 371 (1) | 27.6 | BPD36 | 7 |
| Rindfleisch 1996^170^ | USA | NA | Yes | BPD | 36 (2) | 27.6 | BPD28 | 6 |
| Rocha 2010^171^ | Portugal | 2003 | No | BPD | 205 (1) | 28.6 | BPD36 | 7 |
| Rocha 2011^172^ | Portugal | 2005 | Yes | BPD | 156 (2) | 28.7 | BPD28 | 7 |
| Rojas 1995 ^173^ | USA | 1990 | Yes | BPD | 98 (1) | 26.8 | BPD28 | 9 |
| Ronkainen 2015 ^174^ | Finland | 2000 | Yes | BPD | 88 (1) | 28.7 | BPD28 & BPD36  Mild BPD | 7 |
| Rutkowska 2018 ^175^ | Poland | 2015 | Yes | BPD | 707 (47) | 28.8 | BPD28  Mild/Mod BPD  Sev BPD | 9 |
| Sallmon 2022 ^176^ | Germany | 2017 | Yes | BPD | 34 (1) | 24.6 | BPD-PH | 9 |
| Sampath 2015^177^ | USA | NA | Yes | BPD | 659 (6) | 27.7 | BPD36  Mod BPD  Sev BPD | 6 |
| Sanchez-Soliz 2012 ^178^ | Spain | NA | No | BPD | 75 (1) | 27.9 | BPD28 | 9 |
| Schena 2015^179^ | Italy | 2010 | No | BPD | 242 (1) | 26.3 | BPD28 | 9 |
| Serenius 2004^180^ | Sweden | 1995 | No | BPD | 140 (2) | 24.4 | BPD36 | 8 |
| Sheth 2019 ^181^ | USA | 2014 | No | BPD | 220 (1) | 25.9 | BPD-PH | 9 |
| Shim 2017 ^182^ | Korea | 2014 | No | sex | 1839 (69) | 27.1 | BPD36 | 8 |
| Shinwell 2007 ^183^ | Israel | 1999 | Yes | sex | 8858 (28) | 28.7 | BPD36 | 9 |
| Siddaiah 2022 ^184^ | USA | NA | Yes | BPD | 46 (1) | 25.7 | BPD-PH | 7 |
| Silva da 2018 ^185^ | Brazil | 2016 | Yes | BPD | 67 (3) | 29.1 | BPD36 | 9 |
| Skiöld 2014 ^186^ | Sweden | 2006 | Yes | sex | 107 (NA) | 25.6 | BPD28 & BPD36 | 8 |
| Slaughter 2011 ^187^ | USA | 2006 | No | BPD | 78 (3) | 25.0 | BPD-PH | 8 |
| Smith 2005 ^188^ | USA | 1998 | No | BPD | 5115 (6) | 29.9 | BPD28 & BPD36 | 9 |
| Soliman 2017 ^189^ | Canada | 2009 | Yes | BPD | 319 (1) | 29.0 | BPD36 | 8 |
| Spears 2004 ^190^ | USA | 1996 | No | BPD | 331 (1) | 29.0 | BPD36 | 9 |
| Starr 2020 ^191^ | USA | 2014 | No | BPD | 546 (24) | 27.9 | BPD28 | 9 |
| Steen 2014 ^192^ | Sweden | 2002 | No | sex | 2419 (NA) | 30.0 | BPD36 | 9 |
| Stevenson 2000 ^193^ | USA | 1992 | Yes | sex | 5493 (12) | 28.5 | BPD28 & BPD36 | 7 |
| Stichel 2011^194^ | Sweden | NA | Yes | BPD | 51 (1) | 26.3 | BPD28 & BPD36  Mild BPD | 8 |
| Streimish 2012 ^195^ | USA | 2003 | Yes | BPD | 1103 (14) | 26.5 | BPD36 | 9 |
| Streubel 2008^196^ | USA | 1999 | No | BPD | 133 (1) | 26.3 | BPD28 & BPD36  Mild BPD | 7 |
| Sun 2019 ^197^ | China | 2017 | No | BPD | 296 (1) | 29.8 | BPD28 | 7 |
| Suzuki 2018 ^198^ | Japan | 2012 | Yes | BPD | 45 (1) | 26.5 | BPD36 | 7 |
| Svedenkrans 2019 ^199^ | Australia | 2015 | Yes | BPD | 219 (1) | 27.9 | BPD28 & BPD36  Mild BPD  Mild/Mod BPD  Mod BPD  Sev BPD | 7 |
| Tammela 1992 ^200^ | Finland | 1988 | Yes | BPD | 46 (1) | 31.0 | BPD28 | 7 |
| Tapia 2006 ^201^ | South America | 2002 | Yes | BPD | 1825 (16) | 30.3 | BPD28 | 9 |
| Tauzin 2012 ^202^ | France | 2009 | No | BPD | 137 (1) | 27.8 | BPD36 | 8 |
| Taylor 2014 ^203^ | USA | 2009 | No | BPD | 142 (NA) | 26.2 | BPD36  Mod BPD  Sev BPD | 9 |
| Teberg 1991 ^204^ | USA | 1983 | Yes | BPD | 236 (1) | 30.4 | BPD28 | 9 |
| Tioseco 2006 ^205^ | USA | 1998 | No | sex | 833 (1) | 30.9 | BPD36 | 9 |
| Tokuriki 2015^206^ | Japan | 2013 | Yes | BPD | 25 (1) | 28.9 | BPD36 | 9 |
| Torchin 2016 ^207^ | France | 2011 | Yes | BPD | 2111 (NA) | 29.6 | BPD36 | 9 |
| Uberos 2020 ^208^ | Spain | 2013 | No | BPD | 389 (1) | 29.8 | BPD36 | 9 |
| Van Mastrigt 2018 ^209^ | The Netherlands | 2014 | Yes | BPD | 111 (1) | 28.0 | BPD28 | 9 |
| Vayalthrikkovil 2019 ^210^ | Canada | 2016 | Yes | BPD | 321 (1) | 26.2 | BPD-PH |  |
| Viscardi 2004^211^ | USA | 2001 | Yes | BPD | 262 (2) | 27.6 | BPD28 | 9 |
| Voynow 2020 ^212^ | USA | 2015 | Yes | BPD | 257 (NA) | 26.6 | BPD36 | 9 |
| Vu 2019 ^213^ | Australia | 2013 | No | sex | 430 (1) | 27.9 | BPD36 | 9 |
| Vyas-Read 2018 ^214^ | USA | 2012 | No | BPD | 334 (2) | 26.0 | BPD-PH | 9 |
| Wang 2010 ^215^ | Taiwan | 2004 | Yes | BPD | 72 (2) | 28.3 | BPD28 & BPD36  Mild BPD  Mild/Mod BPD  Mod BPD  Sev BPD | 9 |
| Wang 2013 ^216^ | USA | 2007 | Yes | BPD | 1726 (128) | 26.8 | BPD36 | 9 |
| Wang 2014^217^ | China | 2012 | Yes | BPD | 73 (1) | 30.5 | BPD28 | 7 |
| Wang 2018 ^218^ | Taiwan | 2011 | Yes | BPD | 3507 (20) | 28.0 | BPD36 | 8 |
| Wang 2022 ^219^ | China | 2016 | No | BPD | 268 (1) | 28.2 | BPD-PH | 8 |
| Wang 2023 ^220^ | China | NA | Yes | BPD | 88 (1) | 28.2 | BPD-PH |  |
| Weismann 2017 ^221^ | USA | 2012 | Yes | BPD | 159 (1) | 26.0 | BPD-PH | 9 |
| Wemhöner 2011 ^222^ | Austria | 2005 | No | BPD | 95 (1) | 27.7 | BPD28 | 7 |
| Won 2020 ^223^ | Korea | 2014 | Yes | BPD | 521 (1) | 27.4 | BPD28 | 7 |
| Won Choi 2006 ^224^ | Korea | 2000 | Yes | BPD | 75 (1) | 28.5 | BPD36 | 9 |
| Won Choi 2012 ^225^ | Korea | 2008 | No | BPD | 1180 (77) | 27.5 | BPD36  Mild BPD  Mild/Mod BPD  Mod BPD  Sev BPD | 6 |
| Wu 2013 ^226^ | Taiwan | 2009 | Yes | BPD | 51 (2) | 29.5 | BPD28 | 9 |
| Xie 2016^227^ | China | 2010 | Yes | BPD | 35 (1) | 28.4 | BPD28 | 8 |
| Yilmaz 2017 ^228^ | Turkey | 2016 | Yes | BPD | 40 (1) | 30.2 | BPD28 | 7 |
| Zhang 2011^229^ | China | 2004 | No | BPD | 116 (1) | 30.2 | BPD28 | 7 |
| Zhou 2019 ^230^ | China | 2013 | Yes | BPD | 126 (1) | 28.5 | BPD28 | 7 |
| Zozaya 2019 ^231^ | Spain | 2010 | No | sex | 15125 (NA) | 28.5 | BPD36 | 8 |

BPD: bronchopulmonary dysplasia, GA: gestational age, Mod: moderate, NA: not available; NOS: Newcastle-Ottawa scale, Sev: severe.

**Supplementary Table 2.** Data on heterogeneity of the Bayesian model-averaged meta-analysis of the association between infant sex and any bronchopulmonary dysplasia defined as oxygen requirement during the first 28 days of life or at postnatal day 28.

| **Subgroup** | | **K** | **Heterogeneity (Tau)** | **Credible interval** | | **BFrf** | **Evidence for** | | **frequentist P-value for heterogeneity** |
| --- | --- | --- | --- | --- | --- | --- | --- | --- | --- |
|  |  |  |  | **Lower Limit** | **Upper Limit** |  | **Random effects** | **Fixed effects** |  |
| **All** | | 78 | 0.12 | 0.08 | 0.16 | Inf | Extreme |  | <0.00001 |
| **Mean or median GA** | <27 weeks | 11 | 0.09 | 0.04 | 0.18 | 0.51 |  | Weak | 0.291 |
|  | >27 weeks | 67 | 0.12 | 0.08 | 0.18 | 4.82E+16 | Extreme |  | <0.00001 |
| **Number of infants** | <500 | 60 | 0.10 | 0.05 | 0.17 | 1.47 | Weak |  | 0.013 |
|  | >500 | 18 | 0.13 | 0.08 | 0.20 | 2.47E+19 | Extreme |  | <0.00001 |
| **(sub) Continent** | Eastern Asia | 16 | 0.26 | 0.16 | 0.42 | Inf | Extreme |  | 0.101 |
|  | Europe | 26 | 0.14 | 0.07 | 0.24 | 201328.70 | Extreme |  | 0.00001 |
|  | Latin America | 4 | 0.19 | 0.05 | 0.55 | 0.70 |  | Weak | 0.446 |
|  | North America | 23 | 0.10 | 0.05 | 0.17 | 766.52 | Extreme |  | 0.00030 |
|  | Oceania | 3 | 0.23 | 0.05 | 0.78 | 0.84 |  | Weak | 0.076 |
|  | Western Asia | 6 | 0.24 | 0.09 | 0.51 | 120.43 | Extreme |  | 0.0034 |
| **SDI** | High | 51 | 0.11 | 0.07 | 0.16 | 5.00E+12 | Extreme |  | <0.00001 |
|  | High-Middle | 16 | 0.16 | 0.08 | 0.30 | 23163.74 | Extreme |  | 0.00012 |
|  | MiddIe | 10 | 0.16 | 0.05 | 0.37 | 0.78 |  | Weak | 0.186 |

*GA* gestational age, *SDI* sociodemographic index, *BF* Bayes factor

**Supplementary Table 3.** Data on heterogeneity of the Bayesian model-averaged meta-analysis of the association between infant sex and bronchopulmonary dysplasia defined as oxygen or respiratory support requirement at the post-menstrual age of 36 weeks.

| **Subgroup** | | **K** | **Heterogeneity (Tau)** | **Credible interval** | | **BFrf** | **Evidence for** | | **frequentist P-value for heterogeneity** |
| --- | --- | --- | --- | --- | --- | --- | --- | --- | --- |
|  |  |  |  | Lower Limit | Upper Limit |  | **Random effects** | **Fixed effects** |  |
| **All** | | 123 | 0.09 | 0.07 | 0.12 | 3.02E+17 | Extreme |  | <0.00001 |
| **Mean or median GA** | ≤23 weeks | 2 | 0.14 | 0.05 | 0.37 | 0.45 |  | Weak | 0.731 |
|  | ≤25 weeks | 6 | 0.11 | 0.04 | 0.24 | 0.26 |  | Moderate | 0.237 |
|  | <27 weeks | 44 | 0.07 | 0.04 | 0.11 | 60.02 | Strong |  | 0.015 |
|  | >27 weeks | 77 | 0.10 | 0.06 | 0.15 | 6.93E+07 | Extreme |  | 0.00001 |
| **Number of infants** | <500 | 79 | 0.13 | 0.06 | 0.21 | 9.62 | Moderate |  | 0.069 |
|  | >500 | 42 | 0.09 | 0.06 | 0.12 | 1.16E+15 | Extreme |  | <0.00001 |
| **(sub) continent** | Eastern Asia | 15 | 0.08 | 0.04 | 0.17 | 0.11 |  | Moderate | 0.366 |
|  | Europe | 47 | 0.13 | 0.07 | 0.21 | 1.52E+10 | Extreme |  | <0.00001 |
|  | Latin America | 3 | 0.31 | 0.06 | 1.00 | 1.51 | Weak |  | 0.160 |
|  | North America | 50 | 0.08 | 0.04 | 0.12 | 5.57 | Moderate |  | 0.00080 |
|  | Oceania | 4 | 0.21 | 0.07 | 0.50 | 8.66 | Moderate |  | 0.013 |
|  | Western Asia | 2 | 0.23 | 0.05 | 0.71 | 0.57 |  | Weak | 0.438 |
| **SDI** | High | 100 | 0.07 | 0.05 | 0.11 | 2.27E+08 | Extreme |  | <0.00001 |
|  | High-Middle | 18 | 0.09 | 0.04 | 0.19 | 0.13 |  | Moderate | 0.622 |
|  | MiddIe | 4 | 0.19 | 0.05 | 0.54 | 0.77 |  | Weak | 0.527 |

*GA* gestational age, *SDI* sociodemographic index, *BF* Bayes factor

**Supplementary Table 4.** Data on heterogeneity of the Bayesian model-averaged meta-analysis of the association between infant sex and bronchopulmonary dysplasia-associated pulmonary hypertension

| **Subgroup** | | **K** | **Heterogeneity (Tau)** | **Credible interval** | | **BFrf** | **Evidence for** | | **frequentist P-value for heterogeneity** |
| --- | --- | --- | --- | --- | --- | --- | --- | --- | --- |
|  |  |  |  | **Lower Limit** | **Upper Limit** |  | **Random effects** | **Fixed effects** |  |
| **All** | | 36 | 0.12 | 0.05 | 0.23 | 0.42 |  | Weak | 0.787 |
| **Mean or median GA** | ≤25 weeks | 3 | 0.18 | 0.05 | 0.49 | 0.67 |  | Weak | 0.523 |
|  | <27 weeks | 30 | 0.12 | 0.05 | 0.24 | 0.48 |  | Weak | 0.256 |
|  | >27 weeks | 6 | 0.17 | 0.05 | 0.45 | 0.64 |  | Weak | 0.854 |
| **(sub)Continent** | Eastern Asia | 8 | 0.17 | 0.05 | 0.43 | 0.68 |  | Weak | 0.262 |
|  | Europe | 6 | 0.16 | 0.05 | 0.43 | 0.6 |  | Weak | 0.700 |
|  | North America | 20 | 0.13 | 0.05 | 0.27 | 0.6 |  | Weak | 0.717 |
| **SDI** | High | 32 | 0.12 | 0.05 | 0.24 | 0.48 |  | Weak | 0.755 |
|  | MiddIe | 3 | 0.18 | 0.05 | 0.52 | 0.71 |  | Weak | 0.337 |

*GA* gestational age, *SDI* sociodemographic index, *BF* Bayes factor

**Supplementary Table 5.** Data on heterogeneity of the Bayesian model-averaged meta-analysis of the association between infant sex and severe bronchopulmonary dysplasia.

| **Subgroup** | | **K** | **Heterogeneity (Tau)** | **Credible interval** | | **BFrf** | **Evidence for** | | **frequentist P-value for heterogeneity** |
| --- | --- | --- | --- | --- | --- | --- | --- | --- | --- |
|  |  |  |  | **Lower Limit** | **Upper Limit** |  | **Random effects** | **Fixed effects** |  |
| **All** | | 19 | 0.14 | 0.05 | 0.32 | 0.46 |  | Weak | 0.032 |
|  | ≤25 weeks | 3 | 0.19 | 0.05 | 0.53 | 0.69 |  | Weak | 0.412 |
|  | <27 weeks | 7 | 0.17 | 0.05 | 0.40 | 0.86 |  | Weak | 0.236 |
|  | >27 weeks | 12 | 0.16 | 0.05 | 0.42 | 0.40 |  | Weak | 0.036 |
| **(sub)Continent** | Eastern Asia | 6 | 0.18 | 0.05 | 0.45 | 0.91 |  | Weak | 0.200 |
|  | Europe | 5 | 0.25 | 0.06 | 0.76 | 1.18 | Weak |  | 0.121 |
|  | North America | 6 | 0.19 | 0.06 | 0.43 | 1.58 | Weak |  | 0.194 |
| **SDI** | High | 13 | 0.15 | 0.05 | 0.33 | 0.75 |  | Weak | 0.120 |
|  | High-Middle | 2 | 0.29 | 0.06 | 0.99 | 1.26 | Weak |  | 0.717 |
|  | MiddIe | 2 | 0.25 | 0.05 | 0.93 | 0.89 |  | Weak | 0.107 |

*GA* gestational age, *SDI* sociodemographic index, *BF* Bayes factor

**Supplementary Table 6.** Data on heterogeneity of the Bayesian model-averaged meta-analysis of the association between infant sex and mild, mild/moderate and moderate bronchopulmonary dysplasia.

| **Group** | **K** | **Heterogeneity (Tau)** | **Credible interval** | | **BF_rf_** | **Evidence for** | | **frequentist P-value for heterogeneity** |
| --- | --- | --- | --- | --- | --- | --- | --- | --- |
|  |  |  | **Lower Limit** | **Upper Limit** |  | **Random effects** | **Fixed effects** |  |
| **Mild BPD** | 16 | 0.12 | 0.06 | 0.22 | 3.27E+03 | Extreme |  | 0.021 |
| **Mild/moderate BPD** | 9 | 0.11 | 0.05 | 0.24 | 2.17 | Weak |  | 0.015 |
| **Moderate BPD** | 10 | 0.14 | 0.05 | 0.31 | 1.18 | Weak |  | 0.224 |

*BF* Bayes factor

**Supplementary Table 7.** Robust Bayesian meta-analysis (RoBMA) of the association between infant sex and bronchopulmonary dysplasia.

| Outcome | **K** | **Risk ratio** | **Credible interval** | | **BF_10_** | **BF_rf_** | **BF_bias_** |
| --- | --- | --- | --- | --- | --- | --- | --- |
|  |  |  | **Lower Limit** | **Upper Limit** |  |  |  |
| **BPD28** | 78 | 1.13 | 1.04 | 1.19 | 13.81 | >10^12^ | 0.70 |
| **Mild BPD** | 16 | 1.04 | 0.90 | 1.24 | 0.41 | 188.80 | 0.74 |
| **Mild/Moderate BPD** | 9 | 0.99 | 0.88 | 1.16 | 0.32 | 1.30 | 4.43 |
| **Moderate BPD** | 10 | 1.06 | 0.87 | 1.21 | 0.71 | 1.16 | 1.90 |
| **BPD36** | 123 | 1.21 | 1.15 | 1.26 | >10^4^ | >10^12^ | 1.38 |
| **Severe BPD** | 19 | 1.22 | 1.04 | 1.34 | 8.98 | 0.78 | 1.08 |
| **BPD-PH** | 36 | 0.91 | 0.75 | 1.03 | 0.73 | 0.39 | 0.33 |

*BF* Bayes factor

**Supplementary Table 8.** Bayesian model averaged meta-analysis of the association between infant sex and BPD. Effect size is expressed as odds ratio

| **Outcome** | **Subgroup** | **k** | **Averaged effect (OR)** | **95% credible interval** | | **BF_10_** | **BF_rf_** |
| --- | --- | --- | --- | --- | --- | --- | --- |
|  |  |  |  | **Lower limit** | **Upper limit** |  |  |
| BPD28 | All | 78 | 1.29 | 1.20 | 1.39 | >10^7^ | >10^12^ |
|  | GA<27 weeks | 11 | 1.12 | 1.01 | 1.37 | 2.50 | 0.70 |
|  | GA≥27 weeks | 67 | 1.30 | 1.21 | 1.40 | >10^6^ | >10^9^ |
| BPD36 | All | 123 | 1.37 | 1.32 | 1.42 | >10^23^ | >10^5^ |
|  | GA≤23 weeks | 2 | 1.16 | 0.88 | 1.56 | 1.20 | 0.91 |
|  | GA≤25 weeks | 6 | 1.29 | 1.05 | 1.61 | 7.17 | 8.73 |
|  | GA<27 weeks | 45 | 1.39 | 1.29 | 1.51 | >10^8^ | >10^12^ |
|  | GA≥27 weeks | 78 | 1.39 | 1.32 | 1.46 | >10^14^ | 420.83 |
| Severe BPD | All | 19 | 1.32 | 1.15 | 1.47 | 55.49 | 0.51 |
|  | GA≤25 weeks | 3 | 1.09 | 0.74 | 1.60 | 0.64 | 0.68 |
|  | GA<27 weeks | 7 | 1.19 | 0.93 | 1.53 | 1.07 | 1.16 |
|  | GA≥27 weeks | 12 | 1.34 | 1.16 | 1.51 | 27.83 | 0.39 |
| BPD-PH | All | 39 | 0.92 | 0.81 | 1.04 | 0.47 | 0.29 |
|  | GA≤25 weeks | 3 | 0.89 | 0.58 | 1.33 | 0.73 | 0.68 |
|  | GA<27 weeks | 30 | 0.91 | 0.78 | 1.05 | 0.56 | 0.36 |
|  | GA≥27 weeks | 6 | 1.02 | 0.77 | 1.37 | 0.44 | 0.59 |

*BF* Bayes factor, *BPD* bronchopulmonary dysplasia, *BPD-PH* BPD-associated pulmonary hypertension, *GA* gestational age, *OR* odds ratio. OR>1 indicates male disadvantage.
